# *OSBPL11* is an African-specific locus associated with 25-hydroxyvitamin D concentrations and cardiometabolic health

**DOI:** 10.1101/2025.05.27.25328359

**Authors:** Reagan M Mogire, John Muthii Muriuki, Ruth Fiona Bayimenye, Alexander J Mentzer, Amanda Chong, Mateus H Gouveia, Amy R. Bentley, Gavin Band, Pauline King’ori, Ruth Mitchell, Emily L Webb, Dhriti Sengupta, Lynette Ekunwe, Francis M Ndungu, Alireza Morovat, Alex W Macharia, Clare L. Cutland, Gibran Hemani, Sodiomon B Sirima, Michèle Ramsay, Camila A. Figueiredo, Andrew M. Prentice, Shabir A Madhi, Laura M. Raffield, Manjinder S. Sandhu, Philip Bejon, George Davey Smith, Alison M Elliott, Thomas N Williams, Charles Rotimi, Christina R. Bourne, Anthony Burgett, Adebowale Adeyemo, Sarah H Atkinson

**Author notes:** These authors contributed equally to this work (Adebowale Adeyemo, Sarah H Atkinson).

## Abstract

Vitamin D deficiency is prevalent in Africa, but its genetic determinants are understudied. We report a genome-wide analysis of 25-hydroxyvitamin D (25(OH)D) concentrations in 3670 children from five countries across Africa with replication in four diaspora African ancestry populations (n=21,610). We identify a previously unreported locus at genome-wide significance in West African populations: *OSBPL11* (Oxysterol Binding Protein Like 11, lead variant, rs2979356, p=8.01 × 10^-9^). *In vitro* and molecular docking assays showed that OSBPL11 is a vitamin D binding protein likely involved in the intracellular binding of vitamin D metabolites. *OSBPL11* knockdown mice have increased fat, reduced triglycerides and improved glucose tolerance, and rs2979356 was associated with cardiometabolic health in adults of African ancestry. We also identify previously reported loci determining vitamin D status. Our study advances understanding of vitamin D genetics in Africa and indicates a novel function for *OSBPL11* in intracellular binding and transport of vitamin D metabolites.

## Introduction

Nearly one-third of individuals living in Africa have vitamin D deficiency (25(OH)D of <50 nmol/L), which has been associated with many communicable and non-communicable diseases ^1,2^. Vitamin D is essential for calcium and phosphate homeostasis, skeletal health, and immune function, and its deficiency has been linked to bone disorders, increased risk of infection, metabolic syndrome, and cardiovascular disease^2^. However, the genetics of vitamin D status in Africa remains understudied. Vitamin D genome-wide association studies (GWAS) have been conducted almost entirely in populations of European ancestry^3,4^, while existing knowledge of the genetics of vitamin D status in continental Africans is limited to a few gene-based studies^5,6^. Several vitamin D GWASs have been conducted in individuals of African ancestry in the USA and UK^7–9^, however diaspora African populations have approximately 20% European admixture, and many have different lifestyles and diets compared to continental Africans^10^.

Due to their greater genetic diversity, continental African populations provide a unique platform for investigating the genetic influences on vitamin D status^11^. This genetic diversity increases the likelihood of identifying novel vitamin D loci, which may potentially improve our understanding of vitamin D metabolism and physiology. Furthermore, African populations have relatively smaller linkage disequilibrium blocks, which may improve fine mapping of known loci and facilitate the identification of causal variants and their functional implications in relation to vitamin D status^12^. These unique African genetic characteristics highlight the importance of studying diverse populations to gain a comprehensive understanding of the genetic architecture of vitamin D status.

To our knowledge this is the first GWAS of vitamin D concentrations in continental African populations and includes data from 3,670 children living in Kenya, Uganda, Burkina Faso, The Gambia, and South Africa. We identify a previously unreported genome-wide significant locus associated with vitamin D concentrations, *Oxysterol Binding Protein Like* 11 (*OSBPL11*). Molecular docking and *in vitro* binding assays show that OSBPL11 binds to vitamin D metabolites with comparable conformations and binding energies as common oxysterols, suggesting that OSBPL11 may have a role in intracellular binding and trafficking of vitamin D metabolites. *OSBPL11* knockdown mice have altered glucose tolerance, body fat and triglyceride levels, and the lead *OSBPL11* SNP was associated with cardiometabolic health in African ancestry individuals in continental and diaspora populations.

## Results

### Characteristics of discovery sample

In total, 3670 children with a median age of 23.9 months (interquartile range (IQR) 12.7, 26.0) from Kenya, Uganda, Burkina Faso, The Gambia, and South Africa, had 25(OH)D measurements and genome-wide genotypes (Fig. 1A). Median 25(OH)D concentration was 77.3 nmol/L (IQR 63.5, 93.2). The distribution of 25(OH)D concentrations was right skewed as expected and was rank-based inverse-normal transformed to normalise the distribution (Supplementary Fig. 1). Median body mass index was low and the prevalence of malaria parasitemia and inflammation was high across the cohorts (Supplementary Table 1). The principal component analysis (PCA) plot of genotypes showed clustering of study participants by country and geographical region (Fig. 1B).

**Figure 1.**
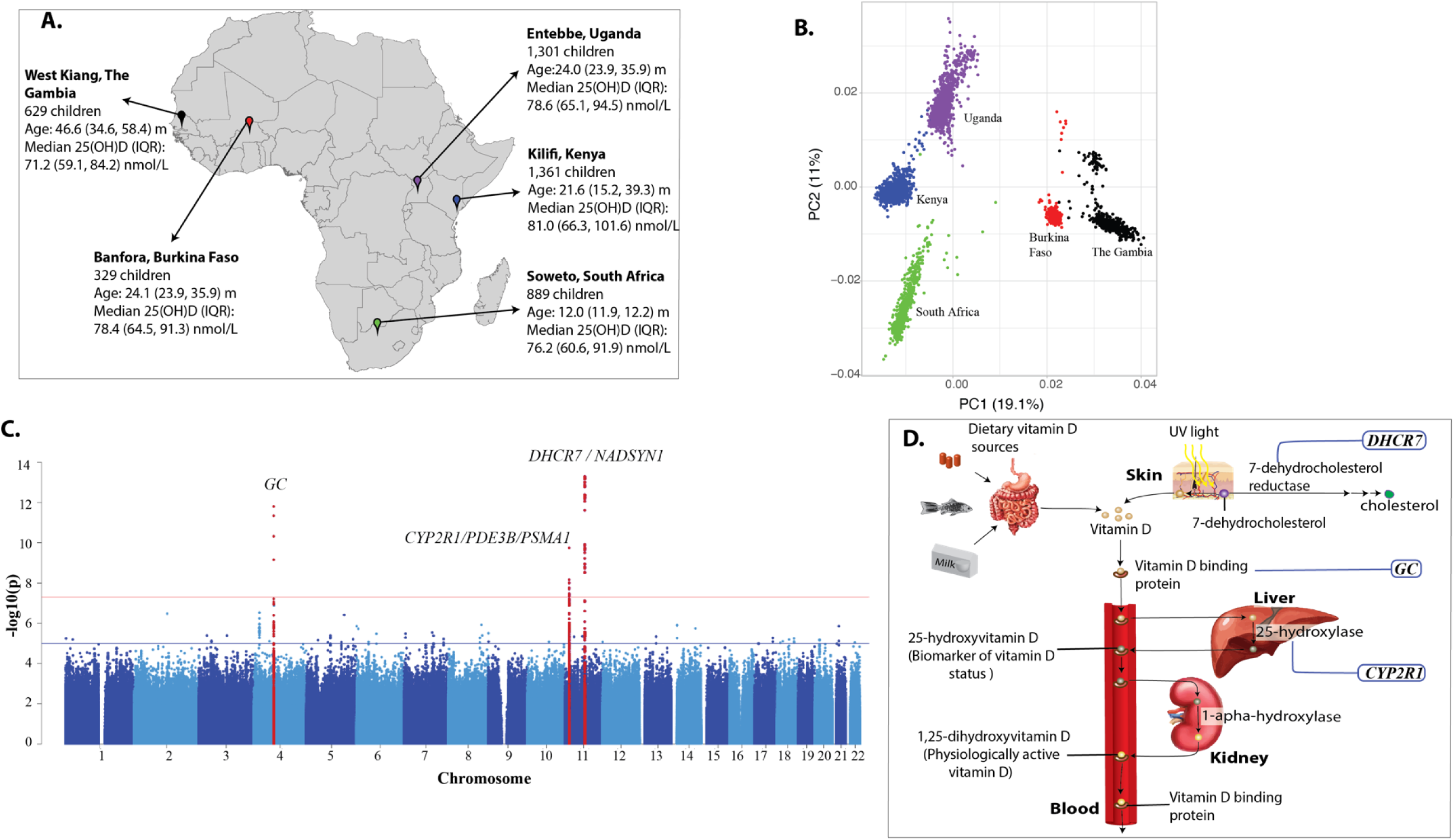
Map of Africa showing locations of discovery cohorts included in the current study (A), principal component analysis (PCA) plot (B), Manhattan plot of discovery GWAS of 25(OH)D concentrations in African children (C), vitamin D metabolism pathway showing effects of genetic loci (D). Abbreviation PC, principal component: IQR, interquartile range. GWAS analyses were adjusted for age, sex, and season of 25(OH)D measurement.

### Discovery genetic association analysis

A total of 18,536,481 variants with minor allele frequency (MAF) > 0.01 were included in the GWAS analyses. Of these, three loci comprising of 61 variants reached genome-wide significance in a meta-analysis of all discovery cohorts (*P* <5×10^−8^). The distribution of the association statistics for the genome-wide association analysis is shown in Fig. 1C. The association analyses were conducted by study site and adjusted for age, sex, and season, and incorporated a Genomic Relationship Matrix (GRM) to account for cryptic relatedness. There was little evidence of inflation of the association analyses (λ=0.99 – 1.01, Supplementary Fig. 2A). The results of the genome-wide association analyses for the independent lead variants are summarized in Table 1.

**Table 1.** Independent genome-wide significant variants for circulating 25(OH)D concentrations in the discovery meta-analysis GWAS for all cohorts (A) and for West Africans (B)

| Gene / Loci | Variant* | Chromosome:<br>position | Effect/ref.<br>allele | Functional<br>effect | Allele<br>Freq. | Effect<br>(Beta) | SE | P value |
| --- | --- | --- | --- | --- | --- | --- | --- | --- |
| **A. All cohorts\*** |  |  |  |  |  |  |  |  |
| *GC* | rs1352846 | 4:72617775 | A/G | intronic | 0.082 | -0.287 | 0.0402 | $1.56 \times 10^{-12}$ |
| *CYP2R1/PDE3B/PSMA1* | rs12290926 | 11:14695046 | A/G | intronic | 0.15 | -0.200 | 0.0313 | $1.77 \times 10^{-10}$ |
| *DHCR7/NADSYN1* | rs7950991 | 11:71158612 | G/T | intronic | 0.17 | 0.220 | 0.0292 | $5.08 \times 10^{-14}$ |
| **B. West Africa** |  |  |  |  |  |  |  |  |
| *OSBPL11* | rs2979356 | 3:125367492 | T/C | intergenic | 0.12 | 0.391 | 0.0678 | $8.01 \times 10^{-9}$ |

\*Overall meta-analysis included cohorts from Kenya, Uganda, Burkina Faso, The Gambia, and South Africa. Additional independent genome-wide significant variants were identified by repeating the associational analyses conditioning for the top variant. No additional signal reached genome-wide significance ( $P$ value of $< 5 \times 10^{-8}$ ) for the three loci.

Three distinct loci, mapped to genes involved in vitamin D metabolism, reached genome-wide significance: the gene for vitamin D binding protein (group-specific component; *GC,* lead SNP rs1352846), vitamin D 25-hydroxylase*/* phosphodiesterase 3B*/* proteasome 20S Subunit Alpha 1 *(CYP2R1/PDE3B/PSMA1*, lead SNP rs12290926) and 7-dehydrocholesterol reductase*/* NAD Synthetase 1 *(DHCR7/NADSYN1*, lead SNP rs7950991) (Fig 1D, Table 1). All the three loci reached genomewide significance in the overall analysis, but only *GC* and *DHCR7/NADSYN1* reached genome-wide significance in the meta-analyses GWAS that included East African, but not West African populations (Supplementary Fig. 2). The widespread distribution of signals across broad genomic regions within the *DHCR7/NADSYN1* locus may suggest selective pressure at this site in continental African populations (Supplementary Fig. 3). The distribution of the association statistics for the individual discovery and replication cohorts are displayed in Supplementary Fig. 4. SNP based heritability for the common variants was 17.6% (standard deviation 8.4%). We found no additional independent loci that reached genome-wide significance in the meta-analysis after performing conditional and joint analysis (GCTA–COJO) on the GWAS summary statistics or after conditioning the individual cohort association analyses for the lead variants in each of the three loci.

### Functional mapping and annotation

Gene-based association analyses showed that the genes that reached genome-wide significance were *GC, CYP2R1, PDE3B, DHCR7,* and *NADSYN1* (Supplementary Fig. 5A, Supplementary Dataset 1). MAGMA gene-set analyses indicated that the top pathways associated with 25(OH)D concentrations were related to intracellular transport to the plasma membrane, vitamin D binding, intracellular signalling, and cell differentiation (Supplementary Dataset 2). Analysis of gene expression data of 53 tissue types from GTEx v8 revealed enriched gene expression in oesophageal mucosa, skin, vagina, and salivary gland tissue (Supplementary Fig. 5B, Supplementary Dataset 3). To predict the most likely causal variants for the three known loci, we performed fine-mapping using FINEMAP (http://christianbenner.com/). *GC, CYP2R1/PDE3B/PSMA1,* and *DHCR7/NADSYN1* were estimated to have 26, 24, and 11 possible causal variants, respectively, with posterior probabilities ranging from 0.54 to 1 (Supplementary Table 2). The variants that were predicted to be most likely to be causal had individual posterior probabilities ranging from 0.40 to 1 (Supplementary Dataset 4). Out of the 60 variants present in the credible sets of the three loci, 26 variants had annotations in PolyPhen (http://genetics.bwh.harvard.edu/pph2/), one was synonymous, and the rest were intronic.

### Meta-analysis with diaspora African populations and transethnic meta-analyses

We performed an African ancestry meta-analysis of vitamin D concentrations that included our discovery GWAS and the Social Change Asthma and Allergy (SCAALA) study in African Brazilians (n=753), African American adults from the Jackson Heart Study (JHS, n=4798) and the Multi-Ethnic Study of Atherosclerosis (MESA, n=1677), adults of African ancestry in the UK Biobank (UKB, n=8151), and precomputed vitamin D GWAS summary data from African ancestry participants in All of Us Research Program (AoS, n=7400)^13^ (Supplementary Table 1). This African ancestry meta-analysis did not reveal any additional genome-wide significant loci (Supplementary Fig. 2B). The three genome-wide significant loci in the discovery GWAS were variably replicated (defined as same direction of effect and *p*<0.05) across the African ancestry cohorts, although direction of effect remained consistent across cohorts (Supplementary Table 3). These loci were also the signals that were most significantly associated with vitamin D status in Europeans. The lead *DHCR7/NADSYN1* variant, rs7950991 was missing in the European GWAS. However, rs12284909, in high linkage disequilibrium with rs7950991 in our discovery cohorts (r^2^=0.98), was genome-wide significant in Europeans (Supplementary Table 3).

### Replication of known 25(OH)D variants in continental Africans

We assessed the transferability of previously reported vitamin D loci in our discovery GWAS. Of 69 independent variants that were genome-wide significant in a recent European vitamin D GWAS ^3^, 66 were present in our discovery GWAS, although with substantial heterogeneity in allele frequency (Supplementary Table 4). Six variants in the *CYP2R1, GC,* SEC23 Homolog A *(SEC23A),* pseudogene *RP13-379L11.3,* and Component of Oligomeric Golgi Complex 5 *(COG5)* genes showed evidence of replication (consistent direction of effect and *P*<0.05). The strongest signals were observed for the *CYP2R1* and *GC* genes. There was little evidence of replication in the remaining 60 variants.

### A novel West African locus in geographical region-specific association analyses

Due to the evidence of population stratification in our discovery cohorts (Fig. 1B), we conducted meta-analyses by geographical region to identify region-specific loci. In a meta-analysis GWAS of West Africans (Burkinabes and Gambians), we identified a novel locus that reached genome-wide significance linked to oxysterol binding protein like 11 (*OSBPL11*). The T allele of variant rs1352846, upstream of *OSBPL11*, was associated with higher circulating 25(OH)D levels (β = 0.391, SE=0.0678, *P*=8.01 × 10^-9^, Fig. 2A). Among West African children, median 25(OH)D levels varied significantly by rs2979356 genotype; 72.9 nmol/L for CC homozygotes, 80.9 nmol/L for the TC heterozygotes, and 90.6 nmol/L for TT homozygotes, demonstrating a strong allele effect (Fig. 2B). *OSBPL11* encodes a member of the oxysterol-binding protein (OSBP) family, a group of intracellular lipid receptors^14^. Regional LocusZoom plots for *OSBPL11* show that the top variants are in high LD and are distributed across a wide genomic region in West African populations (Fig. 2C and Supplementary Fig. 6A, B). FINEMAP predicted a credible set of 8 causal variants (*k=*8) with a posterior probability of 1 (Supplementary Table 2) and the individual variants in the credible set had posterior probabilities ranging from 0.33 to 1 (Supplementary Dataset 4).

**Figure 2.**
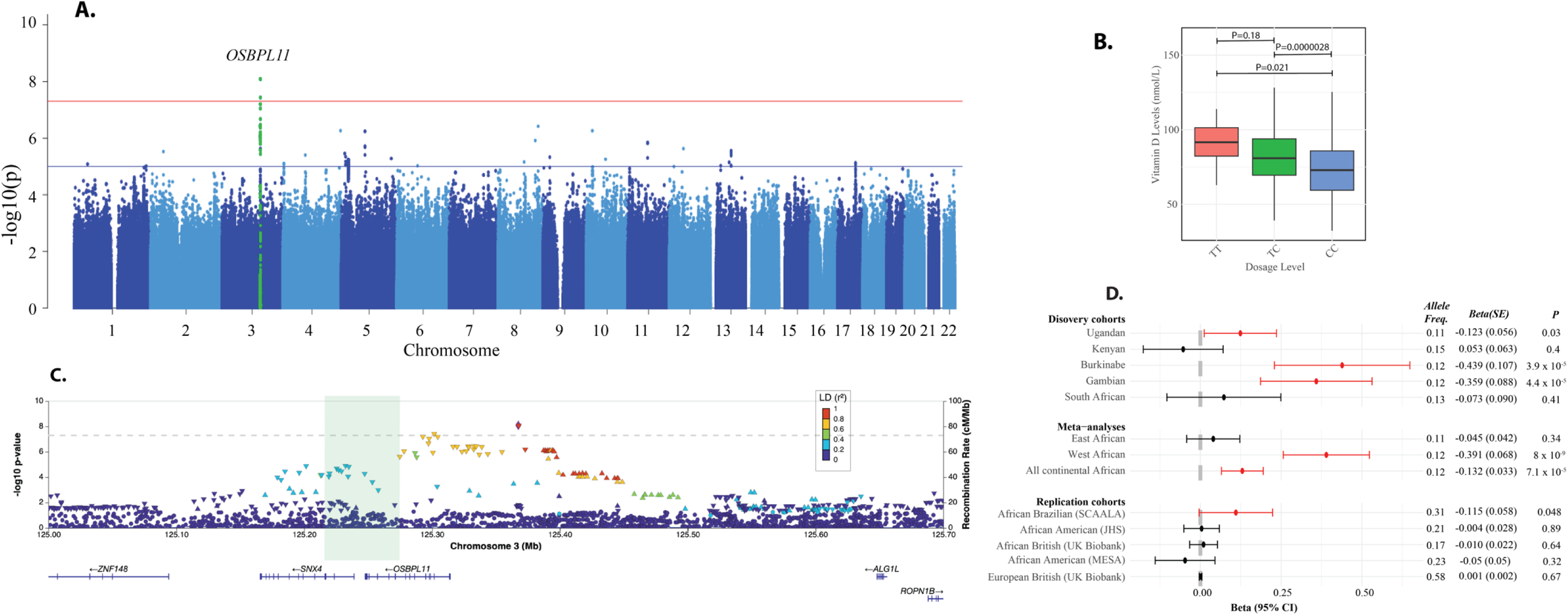
Manhattan plot of meta-analysis GWAS of 25(OH)D concentrations in West African study cohorts (Burkina Faso and The Gambia) (A), boxplot of vitamin D levels by dosages in West African cohorts (B), regional association plot for the *OSBPL11* locus (C), effect of lead variant in the *OSBPL11* locus in discovery and replication cohorts (D), The top variant in the regional plot is shown in a purple diamond shape and the rest are colored according to their LD (R^2^) relative to the top variant in individuals of West African ancestry in this study. The direction of the triangle of the top variant in each locus in the regional plot indicates the direction of effect on 25(OH)D concentrations. Plot B boxplots display the median, interquartile range (IQR), and whiskers extending to 1.5×IQR. Outliers beyond this range were excluded from the plot. Plot D error bars represent 95% confidence intervals (CI), calculated as Effect ± 1.96 × Standard Error (SE).

### Replication and annotation of *OSBPL11*

In addition to Burkinabe and Gambian children, the lead variant in the *OSBPL11* loci, rs2979356, replicated in Ugandan and Brazilian children of African ancestry, but not in adults of African ancestry in the US and the UK (Fig. 2D). Notably, the rs2979356 T allele was the minor allele in African ancestry populations, with a frequency ranging from 0.11 to 0.31, but the major allele in European ancestry populations, where its frequency was 0.58 (Fig. 2D). The nearest signal in the European GWAS is located 219 kilobases away and is marked by the lead variant rs6438900. This variant is mapped to the *MRPL3* (Mitochondrial Ribosomal Protein L3) gene based on genomic proximity (Supplementary Fig. 6). However, rs6438900 and rs2979356 are in weak linkage disequilibrium in Europeans (R²=0.0059) or Gambians (R²=0.0) (LDpair, see Web Resources), suggesting that these signals are distinct. Both rs6438900 and rs2979356 are significant expression quantitative trait loci (eQTLs) for *OSBPL11* in the Open Targets Genetics database, but only rs2979356 is a significant eQTL for *OSBPL11* in the African Functional Genomic Resource and the JHS eQTL Database (see Web Resources). However, the rs2979356 is not an eQTL lead variant or part of an established eQTL credible set in the Open Targets Genetics database. We found no evidence of epistasis between these variants in the Burkinabe and Gambian cohorts (*P* > 0.05), suggesting no combined effect on the phenotype.

### *In vitro* and molecular docking show vitamin D metabolites are ligands of OSBPL11

Due to the strong similarity in molecular structure of vitamin D metabolites and oxysterols, we hypothesized that they might share binding proteins such as OSBPL11 (Fig. 3A). However, OSBPL11 protein structures and *in vitro* ligand binding studies have not previously been reported. The AlphaFold model of the OSBPL11 ligand binding domain shows broad similarities to existing OSBP and OSBPL2 protein structures^15,16^ and similarly adopts a canonical incomplete beta barrel capped by a likely flexible “lid” loop under which ligands are accommodated (arrow) (Fig. 3B and Supplementary Fig. 7). Docking scores support a high potential to interact with both oxysterols and vitamin D compounds. The side-chain hydroxylated oxysterols were all shown to have only one top scoring docked pose, indicating more constrained binding orientations compared to the vitamin D compounds (Fig. 3C). Importantly, both 25-OHC and calcitriol interact with low nanomolar affinity (inhibition constants [Ki] of 32 nM and 64 nM, respectively) in 293T lysate overexpressing full length OSBPL11-myc-His, using a well-established ^3^H-25-OHC competition binding assay (Fig. 3D)^17–19^. These results, which are the first reported *in vitro* ligand binding results of OSBPL11, clearly support OSBPL11 as a vitamin D binding protein and imply that OSBPL11 may be involved in the intracellular transport of vitamin D metabolites (Fig. 3E).

**Figure 3.**
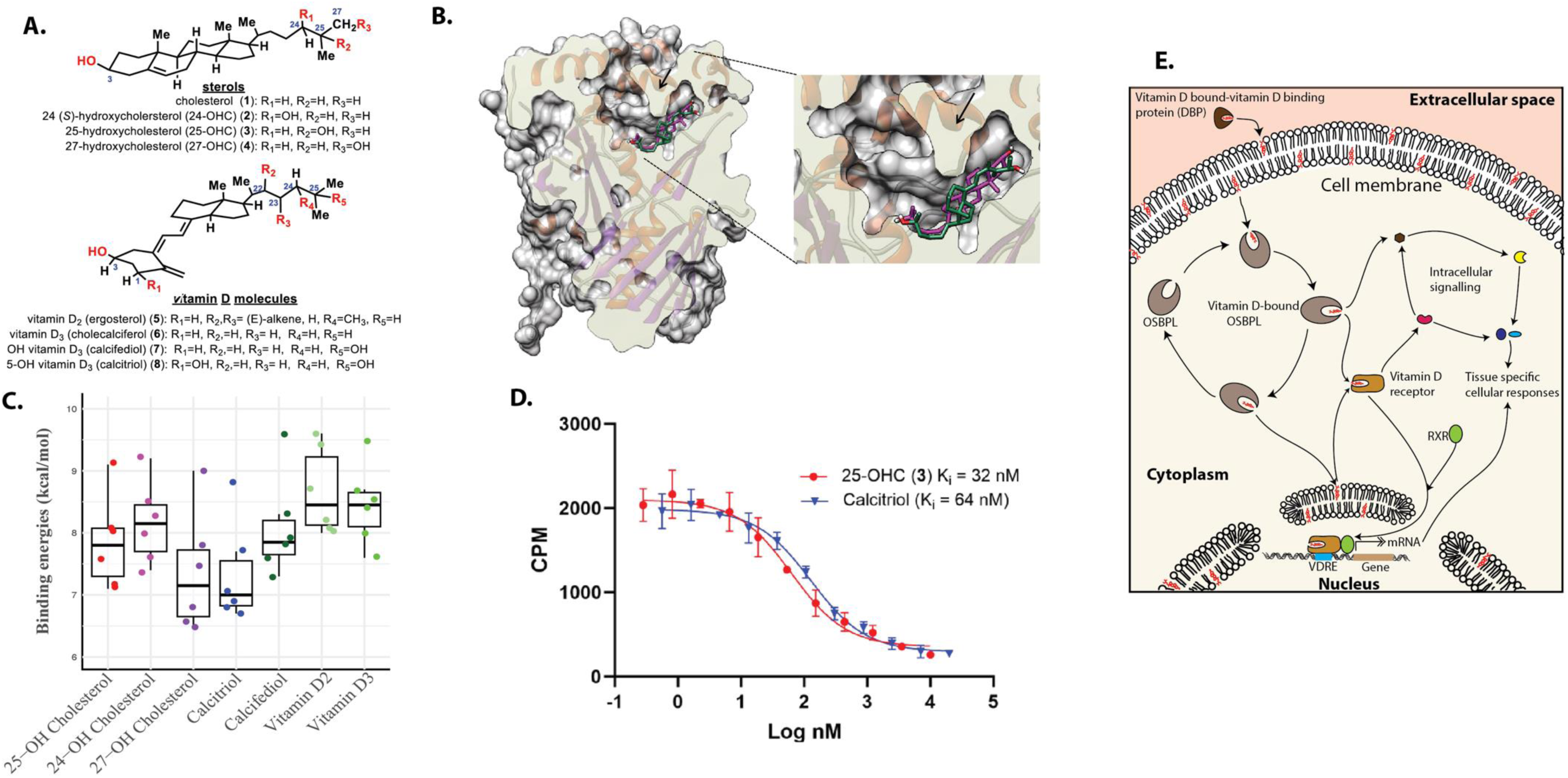
Molecular structures of sterols and vitamin D compounds (A), molecular docking of 25-hydroxycholesterol (magenta) and calcitriol (1,25-OH vitamin D3) (green) into Alpha Fold modeled OSBPL11 (Q9BXB4 v4) (B), Top six docking scores of common oxysterols and vitamin D metabolites on OSBPL11 Alpha Fold model (C), ^3^H-25-hydroxycholesterol-competition binding curves of 25-hydroxycholesterol (25-OHC) and calcitriol on OSBPL11 in *in vitro* OSBP/ORP competition binding assay (D), and illustration showing hypothesized mechanism of action of OSBPL11 in vitamin D mediated physiological activities (F). *In vitro* binding was performed on overexpressed OSBPL11-myc-His, in S100 293T lysate, using established 3H-25-OHC *in vitro* OSBP/ORP competition binding assay. Past studies suggest OSBPL11 may be involved in shuttling 1,25-dihydroxyvitamin D between intracellular membranes and ligand receptors (adapted from ^14^).

### *In silico* functional follow up of *OSBPL11*

*OSBPL11* is expressed in many tissues and cell types (Supplementary Fig. 8A and B) and eQTL summary statistics for rs2979356 indicate that it is a significant eQTL for multiple genes across various tissues, suggesting its widespread regulatory impact on gene expression (Supplementary Dataset 5). *OSPL11* is most highly associated with increased expression in adipocytes, whole blood and monocytes (Supplementary Fig. 9A), and differential expression is also associated with various glucose metabolism and obesity related clinical phenotypes and cardiovascular diseases and obesity (Supplementary Fig. 9B and C, Supplementary Dataset 6). Single cell atlas of human and mouse white adipose tissue reveal that *OSBPL11* is highly expressed in adipocytes, macrophages, endothelial, and circulating mononuclear cells (Supplementary Fig. 10), and rs2979356 is located in super enhancers, chromatin regulatory elements and DNA methylation sites in adipose and other tissues (Haploreg, see Web Resources for links). PheWAS data linked rs2979356 to several health traits, including OSBPL11 levels (Supplementary Dataset 7, https://gwas.mrcieu.ac.uk/phewas). The STRING database shows that OSBPL11 binds to 25-hydroxycholesterol and is involved in fatty acid metabolism and regulating adipocyte triglyceride storage (https://string-db.org/).

Given the suggestive evidence of selection pressure in the region surrounding *OSBPL11* and the role of OSBP in modulating inflammatory responses^20^ we investigated whether this might be driven by protection from severe malaria or tuberculosis (TB), but found little evidence that the rs2979356 variant affects risk of severe malaria (β(SE) = -0.0396 (0.032), *P=*0.21)^21^ or tuberculosis infection (odds ratio (SE) = 0.993 (0.054), *P=*0.91)^22^ in continental Africans. As of December 2024, there were no data available for rs2979356 in the NHGRI-EBI GWAS catalog, PheGenI, dbGaP, ClinVar, PopHumanScan or in EGA (see Web Resources for links).

### *In vivo* mouse studies on *OSBPL11* knockouts

Heterozygous knockout of *OSBPL11* in mice leads to improved glucose tolerance and increased total body fat with lower circulating triglycerides compared to wild types (*P<*0.0001; International Mouse Phenotyping Consortium–see Web Resources; Supplementary Fig. 11A and B). Other measured phenotypes in heterozygous knockouts showing suggestive evidence (*P<*0.05), include increased fat mass (*P=*0.0017), reduced lean/body weight ratio (*P=*0.0029), and increased bone mineral density (*P=*0.0087). Homozygous *OSBPL11* knockout mice experience preweaning lethality suggesting that OSBPL11 is essential for early postnatal survival and possibly plays a critical role in developmental processes.

### Effect of lead *OSBPL11* variant on cardiometabolic traits in human studies

Considering the association between *OSBPL11* and glucose tolerance, adiposity and triglycerides in mice, we analyzed the effect of rs2979356, the lead variant in this locus, on cardiometabolic traits in continental and diaspora African ancestry populations. We found that rs2979356 was associated with reduced BMI and fat mass in the AADM cohort, as well as with reduced triglycerides in the GLGC and AMP-CMDKP studies and reduced LDL cholesterol in the AWI-Gen study (Table 2). Rs2979356 was also associated with higher risk of hypertension and stroke in the AMP-CMDKP study, and with stroke alone in African ancestry adults in the UKBB. Findings varied between cohorts which might reflect sample size, ancestry or haplotype distributions among populations. We found little evidence of association between rs2979356 and HDL or total cholesterol levels or with fasting glucose or insulin levels (Supplementary Table 5).

**Table 2.** Effect of *OSBPL11* lead variant, rs2979356, on select biomarkers of lipid metabolism and cardiovascular health in African ancestry individuals.

|  | n | Beta | SE | P |
| --- | --- | --- | --- | --- |
| **Triglycerides** |  |  |  |  |
| AADM (overall) | 4,292 | 0.0203 | 0.0318 | 0.52 |
| AADM (West Africa) | 3,565 | -0.00577 | 0.0365 | 0.87 |
| UKB (African ancestry) | 6,211 | -0.0268 | 0.0245 | 0.56 |
| AMP-CMDKP | 128,439 | -0.0155 | 0.0039 | 0.0042 |
| AWI-Gen (overall) | 10,603 | -0.011 | 0.020 | 0.58 |
| AWI-Gen (West Africa) | 3,763 | -0.027 | 0.034 | 0.43 |
| GLGC | 89,467 | -0.0158 | 0.00598 | 0.0082 |
| **LDL cholesterol** |  |  |  |  |
| AADM (overall) | 4,257 | 0.0186 | 0.0322 | 0.56 |
| AADM (West Africa) | 3,550 | 0.0223 | 0.0364 | 0.54 |
| AWI-Gen (overall) | 10,603 | -0.044 | 0.020 | 0.015 |
| AWI-Gen (West Africa) | 3,763 | -0.047 | 0.033 | 0.16 |
| GLGC | 87,759 | -0.00619 | 0.00606 | 0.31 |
| UKB (African ancestry) | 6,006 | 0.0270 | 0.0254 | 0.54 |
| AMP-CMDKP | 115,939 | 0.0055 | 0.0059 | 0.35 |
| **BMI** |  |  |  |  |
| AADM (overall) | 5,454 | -0.0649 | 0.155 | 0.000032 |
| AADM (West Africa) | 4,695 | -0.0186 | 0.175 | 0.0000024 |
| AWI-Gen (overall) | 10,603 | 0.012 | 0.017 | 0.49 |
| AWI-Gen (West Africa) | 3,762 | 0.024 | 0.025 | 0.34 |
| UKB (African ancestry) | 6,545 | -0.0631 | 0.0449 | 0.80 |
| **Fat mass** |  |  |  |  |
| AADM (overall) | 5,295 | -0.158 | 0.302 | 0.000051 |
| AADM (West Africa) | 4,537 | -0.0153 | 0.336 | 0.00000046 |
| **Hypertension** |  |  |  |  |
| AWI-Gen (overall) | 10,708 | 0.010 | 0.009 | 0.27 |
| UKB (African ancestry) | 6,626 | -0.0525 | 0.137 | 0.15 |
| AMP-CMDKP | 31,968 | 0.0512 | 0.0247 | 0.039 |
| **Stroke** |  |  |  |  |
| UKB (African ancestry) | 6,520 | -0.00623 | 0.117 | 0.019 |
| AMP-CMDKP | 4,963 | 0.482 | 0.233 | 0.038 |

## Discussion

The high genetic heritability and environmental susceptibility of vitamin D status underscore the importance of studying it across diverse global populations. This is especially relevant in sub-Saharan Africa due to immense genetic diversity, varied lifestyles and environmental conditions, together with a high risk of communicable and non-communicable diseases. In the present study, we report a genome-wide analysis of circulating 25(OH)D concentrations in 3670 children from five countries across Africa. Our study identified three known genome-wide significant loci in genes directly involved in vitamin D metabolism in overall meta-analysis. We also found a previously unreported locus (*OSBPL11)* for 25(OH)D concentrations. The lead *OSBPL11* variant had a large effect on 25(OH)D concentrations in West African children and was also replicated in Ugandan and Brazilian children of African ancestry. Our *in vitro* binding assays revealed that calcitriol binds to OSBPL11 with similar conformation and binding energies as its known ligands, the common oxysterols. Knockdown of *OSBPL11* in mice improved glucose tolerance, increased total body fat and lowered circulating triglyceride levels. The lead variant in the *OSBPL11* loci was also associated with triglyceride levels, LDL cholesterol, fat mass, BMI, hypertension and stroke in African ancestry populations.

The genome-wide significant signals identified in our overall meta-analysis are located within previously established vitamin D metabolizing genes. The *GC* gene encodes the vitamin D binding protein, responsible for the binding and circulation of 80-90% of vitamin D and its metabolites ^23^. The *GC* locus is the sole locus that reached genome-wide significance in recent GWASs of African American participants^7^ ^24^, and was one of the strongest signals in GWASs in European^3,4^, Hispanic^25^, Asian ^26^ and mixed ancestry^8,27^ populations, indicating it is a multi-ancestry signal. Epidemiological studies in sub-Saharan Africa show strong associations between various DBP isoforms and vitamin D status^5,6^.

Gene-based association analysis revealed that the *CYP2R1* locus had the strongest effect on 25(OH)D concentrations of all genes, likely due to its encoding of the enzyme 25- hydroxylase, which converts vitamin D to 25-hydroxyvitamin D, the biomarker generally used in GWAS of vitamin D status ^28^. The *DHCR7* gene encodes 7-dehydrocholesterol reductase which diverts 7-dehydrocholesterol towards cholesterol synthesis and away from vitamin D synthesis in the skin ^28^. However, the locus was not genome-wide significant in the replication cohort of diaspora African-ancestry populations, possibly because these populations rely less on sun exposure for vitamin D synthesis and more on vitamin D supplementation and food fortification. Although the three loci are replicated in previous European GWASs, we found that only a small proportion (fewer than 10%) of variants that influence 25(OH)D concentrations in European populations replicate in continental African populations. These differences may be due to genetic heterogeneity due to different causal variants (which is common in complex traits), differences in genetic linkage and LD structure, gene-environment interactions, allele frequency differences or the limitations of sample size of our study ^29^.

Our study identified a novel locus, *OSBPL11*, which was associated with higher 25(OH)D concentrations in West African children with replication in Ugandan and Brazilian children of African ancestry. This effect was not observed in Kenyan or South African children, or adults of African and European ancestry (despite the allele being at a higher frequency in the latter), highlighting both subcontinental heterogeneity within African populations and inter-ancestry differences across the study cohorts. A plausible explanation is that the *OSBPL11* locus exhibits different linkage disequilibrium (LD) patterns across Africa and in African versus European ancestry groups leading to ancestry-specific effects. Furthermore, environmental factors, such as dietary differences, sun exposure levels, or lifestyle, could modulate gene–environment interactions in ways that differ across populations.

Our *in vitro* binding assays and molecular docking findings suggest that OSBPL11 interacts with vitamin D compounds, potentially mediating its physiological roles such as intracellular transport of vitamin D metabolites. Unlike other loci in the overall meta-analysis, *OSBPL11* has not previously been reported to be directly linked to vitamin D status or metabolism in the published literature. Studies have shown that lithocholic acid, an oxysterol, can activate vitamin D receptors (VDRs), the same receptors activated by 1,25-dihydroxyvitamin D ^30^, indicating a potential indirect link between vitamin D and oxysterol binding proteins.

Furthermore, both vitamin D and oxysterols are implicated in lipid metabolism and demonstrate similar anti-inflammatory and immunomodulatory effects, suggesting the possibility of shared or intersecting biological pathways^28,31^. OSBPL11, which regulates adipocyte triglyceride storage^32^ and is strongly associated with obesity and overnutrition, may play a role in vitamin D physiology, as higher rates of obesity have been associated with vitamin D deficiency ^2^. Given that OSBPL11 and other members of the OSBP family are implicated in the intracellular transport of lipids^14^, this finding suggests that vitamin D compounds might influence lipid distribution and signaling pathways within cells. This study lays the groundwork for further exploration into how alterations in OSBPL11 function may affect vitamin D homeostasis and contribute to clinical outcomes. For instance, dysregulation of vitamin D signaling has been associated with diseases such as osteoporosis and certain cancers^1,33^, and may explain why obese individuals are on average 35% more likely to be vitamin D deficient than non-obese individuals^34^. Moreover, the response to vitamin D supplementation in African American participants has been linked to methylation in OSPL6, a protein that, like OSBPL11, belongs to the OSBPL family protein ^35^.

*In vivo* and *in vitro* studies collectively highlight the possible roles OSBPL11 plays in lipid and glucose metabolism and metabolic health. The high expression of OSBPL11 in adipocytes and its direct involvement in regulating fatty acid metabolism *in vitro* — specifically through the modulation of proteins such as ADIPOQ and FABP4, as well as adipocyte triglyceride storage, suggest that OSBPL11 is crucial for adipocyte function and lipid homeostasis (STRING database). Mouse studies corroborate these findings, showing that OSBPL11 influences glucose homeostasis, triglyceride levels, and total body fat. In humans, elevated expression of OSBPL11 in adipocytes has been associated with conditions related to overnutrition, such as obesity and other diseases of metabolism. Notably, we found that the lead variant at the *OSBPL11* locus, rs2979356, was linked to increased fat mass and triglyceride levels, as well as heightened risks of stroke and hypertension in African ancestry populations although findings varied across populations (Table 2), suggesting that OSBPL11 may contribute to the predisposition to metabolic disorders prevalent in these populations.

*OSBPL11* gene polymorphisms have been associated with diastolic blood pressure, hyperglycemia/diabetes, low density lipoproteins-cholesterol (LDL) levels and metabolic syndromes in obese individuals^36^. Our study extends the understanding of OSBPL11 by revealing its association with 25-hydroxyvitamin D levels and indicating that OSBPL11 may also influence vitamin D metabolism and transport. The shared pathways between lipid and vitamin D metabolism may explain the observed genetic associations and the overlapping physiological roles of vitamin D and oxysterols^14^, thereby underscoring the multifaceted role of OSBPL11 in metabolic regulation. Further research is warranted to elucidate the molecular mechanisms underlying OSBPL11’s functions and to explore its potential as a therapeutic target for improving metabolic health and reducing disease risk.

The strengths of this study include being the first vitamin D GWAS to be conducted in continental African populations, including community-based children from five countries across Africa. A major highlight of the study is the identification of *OSBPL11*, a novel locus associated with 25(OH)D concentrations, marking the first evidence of its role in vitamin D status. We are also the first study, to the best of our knowledge, to demonstrate that OSBPL11 binds to vitamin D metabolites in molecular docking or *in vitro* analyses. These findings provide critical insights into the potential role of OSBPL11 in intracellular vitamin D transport and metabolism. However, our findings should be interpreted in the context of some limitations. First, our GWAS may not have captured low-frequency variants or variants with small effect sizes since vitamin D is a complex trait that is regulated by many genetic and environmental factors. The relatively smaller sample size of our study compared to the much larger European vitamin D GWAS may partially explain our inability to replicate some of their loci. Additionally, the generalisability of our findings to populations with different ethnicities, age groups, dietary habits, sun exposure and other environmental factors remains uncertain. Moreover, many of the individuals in the African diaspora populations included in our replication cohorts were overweight, middle aged and on vitamin D supplements and thus not very representative of African children.

In summary, our study described the genetic determinants of vitamin D concentrations in individuals of African ancestry. We identified a novel association between vitamin D concentrations and the *OSBPL11* gene, marking a significant advance in understanding the mechanisms underlying vitamin D metabolism. This discovery opens new avenues for investigating how OSBPL11 may influence intracellular vitamin D pathways, particularly its potential role in the transport of vitamin D metabolites within cells. Moreover, the association between the lead variant in *OSBPL11* and cardiometabolic traits, such as triglyceride levels, fat mass, BMI, hypertension, and stroke, suggests a role of OSBPL11 in lipid metabolism.

This finding underscores the possibility that shared pathways between lipid and vitamin D metabolism could contribute to the observed associations. Additionally, our study confirmed previously reported associations between vitamin D concentrations and well-established metabolism genes, namely, *GC, CYP2R1,* and *NADSYN1/DHCR7,* further validating their role in vitamin D physiology. Nevertheless, the lack of transferability of some loci–such as the *OSBPL11*–even within some African populations raises important questions about the generalizability of genetic research findings across diverse ancestry groups and emphasizes the need for greater inclusion of underrepresented populations in genomic studies.

## Supplementary Appendix

**Supplementary Table 1.** Characteristics of study participants.

|  | Discovery cohorts |  |  |  |  | Replication cohorts |  |  |  |  |  |
| --- | --- | --- | --- | --- | --- | --- | --- | --- | --- | --- | --- |
|  | Kenyans | Ugandans | Burkinabe | Gambians | South Africans |  | African<br>Brazilian<br>(SCAALA) | African American<br>Adults from<br>JHS | Multi-Ethnic<br>Study of<br>Atherosclerosis<br>(MESA) | African ancestry<br>UK (UKB) | African American<br>(All of US) |
| No. of<br>participants<sup>§</sup> | 962 | 1288 | 329 | 484 | 607 |  | 753 | 5306 | 1677 | 8151 | 7400 |
| Median 25(OH)D<br>nmol/L (IQR)<sup>*</sup> | 78.1 (64.0, 96.5) | 78.6 (65.0, 94.6) | 78.6 (65.1, 94.5) | 72.4 (59.6, 84.9) | 72.7 (58.7, 88.8) |  | 27.4 (21.3, 33.1) | 32.4 (22.5, 46.2) | 17.4 (12.1, 24.2) | 31.0 (22.1, 42.9) | 26.0 (17.0, 37.0) |
| Median (IQR) age<sup>†</sup> | 21.6 (15.2, 39.3)<br>m | 24.0 (23.9, 35.9)<br>m | 24.1 (23.9, 35.9)<br>m | 46.6 (34.6, 58.4)<br>m | 12.0 (11.9, 12.2)<br>m |  | 7.0 (6.0, 9.0) y | 55.0 (45.0, 64.0) y | 62.0 (53.0, 70.0) | 50.0 (45.0, 58.0) y | 61.0 (50.0, 69.0) y |
| Sex: females | 473/962 (49.2%) | 638/1288 (49.5%) | 161/329 (48.9%) | 218/484 (45.0%) | 291/607 (47.9%) |  | 361/750 (48.1%) | 3367/5306 (63.1%) | 906 (54.0%) | 3393/8151 (41.6%) | 6458/8935 (72.28%) |
| Season<sup>#</sup> |  |  |  |  |  |  |  |  |  |  |  |
| 1<sup>st</sup> season | 48/962 (5.0%) | 302/1284 (23.4%) | 72/329 (21.9%) | - | 238/607 (39.2%) |  | 179 (24.2%) | 1137/5306 (21.4%) | 409/1677(24.4%) | 1719/8151 (21.1%) | 2255/8935 (25.2%) |
| 2<sup>nd</sup> season | 357/962 (37.1%) | 310/1284 (24.1%) | 123/329 (37.4%) | - | 25/607 (4.1%) |  | - | 1189/5306 (22.4%) | 565/1677(33.6%) | 2197/8151 (27.0%) | 1889/8935 (21.1%) |
| 3<sup>rd</sup> season | 528/962 (54.9%) | 318/1284 (24.6%) | 129/329 (39.2%) | 536/484 (85.4%) | 228/607 (37.6%) |  | - | 1447/5306 (27.3%) | 339/1677(20.2%) | 1565/8151 (19.2%) | 2220/8935 (24.9%) |
| 4<sup>th</sup> season | 29/962 (3.0%) | 354/1284 (27.5%) | 5/329 (1.5%) | 92/484 (14.7%) | 116/607 (19.1%) |  | 562 (75.8%) | 1533/5306 (28.9%) | 364/1677(21.7%) | 2670/8151 (32.7%) | 2571/8935 (28.8%) |
| BMI (IQR) | 14.9 (13.7, 16.) | 15.7 (14.7, 16.4) | 15.6 (14.7, 16.4) | 14.6 (13.8, 15.4) | n/a |  | 15.6 (14.3, 16.3) | 30.5 (26.9, 35.4) | 29.4 (26.1, 33.5) | 28.7 (25.7, 32.3) | 32.1 (27.1, 38.4) |
| Malaria<sup>¶</sup> | 189/857 (22.1%) | 88/1268 (6.9%) | 64/303 (21.1%) | 65/484 (10.4%) | <sup>¶</sup>No malaria |  | <sup>¶</sup>No malaria | <sup>¶</sup>No malaria | <sup>¶</sup>No malaria | <sup>¶</sup>No malaria | <sup>¶</sup>No malaria |
| Inflammation<sup>‡</sup> | 256/962 (27.0%) | 304/1273 (23.9%) | 109/329 (33.9%) | 85/484 (14%) | 94/607 (15.5%) |  | 230/751(30.5%) | 1242/4791(25.9%)<sup>§§</sup> | 445/1677 (26.6%) | 1088/8151 (13%) | n/a |
| Supplementation<sup>††</sup> | n/a<sup>††</sup> | n/a<sup>††</sup> | n/a<sup>††</sup> | n/a<sup>††</sup> | n/a<sup>††</sup> |  | n/a<sup>††</sup> | 1648/5306 (34.4%) | n/a | 490/7611 (6.4%) | 5640/8935 (63.1%) |
| Abbreviations; IQR, interquartile range; n/a, not available; 25(OH)D, 25-hydroxyvitamin D. <sup>§</sup>The number of participants in each study cohorts included in the GWAS, based on availability of genotype and phenotype data. <sup>¶</sup>Malaria was defined as the presence of *P. falciparum* parasites on blood film. <sup>‡</sup>Inflammation was defined as CRP > 5 mg/L or ACT > 0.6 g/L. ACT, but not CRP, was available for The Gambia. <sup>†</sup>Age is presented in months for discovery cohorts and years for replication cohorts. <sup>#</sup>1<sup>st</sup> season corresponds to summer/short rains/dry season; 2<sup>nd</sup> season as autumn/dry season; 3<sup>rd</sup> season as winter/long rains season; 4<sup>th</sup> season as spring or dry season. Seasons in discovery cohorts were based on 3 monthly intervals as follows 1<sup>st</sup> season, December to February; 2<sup>nd</sup> season, March to May; 3<sup>rd</sup> season, June to August; 4<sup>th</sup> season, September to November. In South Africa these seasons correspond to summer, autumn, winter, and spring, respectively, in Uganda and Kenya there are two rainy and two dry seasons and in Burkina Faso and The Gambia there is a single rainy and dry season. For MESA study, seasons were defined as follows;1<sup>st</sup> season from January to March; 2<sup>nd</sup>, April to June; 3<sup>rd</sup>, July to September; 4<sup>th</sup>, October to December. However, the timing of the rains is often unpredictable and may vary from these times. <sup>§§</sup>HsCRP was measured in African American cohorts, which was converted to values equivalent to CRP as previously (Arthroplasty 2014). <sup>††</sup>Vitamin D supplementation is very low/non-existent in the African children. |  |  |  |  |  |  |  |  |  |  |  |

**Supplementary Table 2.** Fine-mapping posterior probabilities of *k* causal variants at independent signal loci that reached genome-wide significance in the discovery GWAS.

| Locus | GWAS index variant | Chromosome: position | K (no. of causal variants) * | Posterior probability |
| --- | --- | --- | --- | --- |
| *GC* | rs1352846 | 4:72617775 | **26** | **1** |
| | | | 27 | $2.96 \times 10^{-10}$ |
| *CYP2R1/PDE3B/PSMA1* | rs12290926 | 11:14695046 | **24** | **0.54** |
|  |  |  | 25 | 0.30 |
|  |  |  | 21 | 0.056 |
|  |  |  | 22 | 0.040 |
|  |  |  | 23 | 0.046 |
|  |  |  | 26 | 0.021 |
| *DHCR7/NADSYN1* | rs7950991 | 11:71158612 | **11** | **1** |
| | | | 10 | $2.96 \times 10^{-106}$ |
| *OSBPL11* | rs2979356 | 3:125367492 | **8** | **1** |

\*K (causal variants) 0 to 100 were analyzed. However, only *ks* with non-zero posterior probabilities are presented in this table. The most probable no of causal variants (k) is highlighted in bold. The *k* variants listed are predicted as possible credible sets and are not definitively causal.

**Supplementary Table 3.**
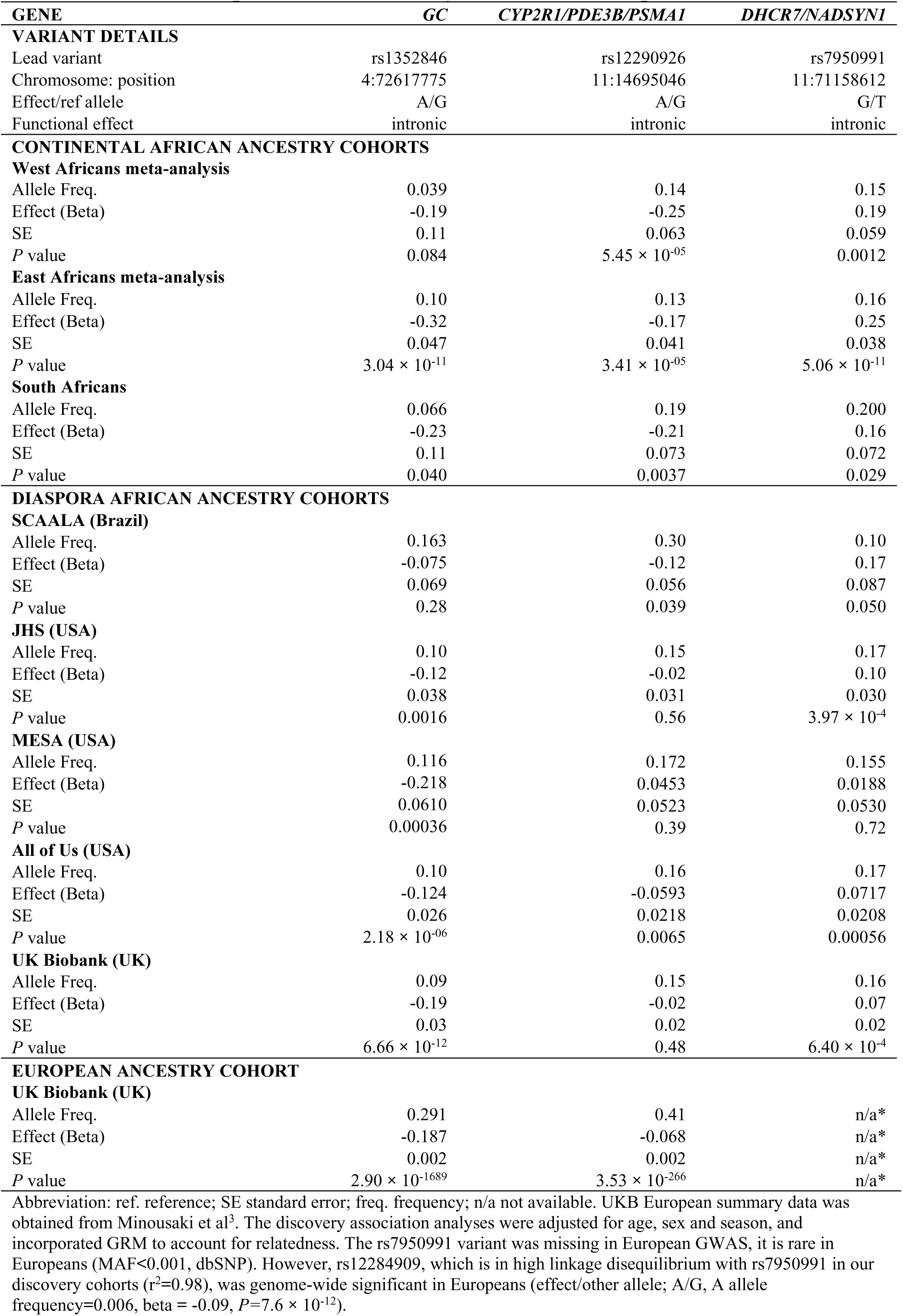
Effect of independent African genome-wide significant variants in continental and diaspora African ancestry cohorts and Europeans in the UK Biobank.

| GENE | GC | CYP2R1/PDE3B/PSMA1 | DHCR7/NADSYN1 |
| --- | --- | --- | --- |
| **VARIANT DETAILS** |  |  |  |
| Lead variant | rs1352846 | rs12290926 | rs7950991 |
| Chromosome: position | 4:72617775 | 11:14695046 | 11:71158612 |
| Effect/ref allele | A/G | A/G | G/T |
| Functional effect | intronic | intronic | intronic |
| **CONTINENTAL AFRICAN ANCESTRY COHORTS** |  |  |  |
| **West Africans meta-analysis** |  |  |  |
| Allele Freq. | 0.039 | 0.14 | 0.15 |
| Effect (Beta) | -0.19 | -0.25 | 0.19 |
| SE | 0.11 | 0.063 | 0.059 |
| P value | 0.084 | $5.45 \times 10^{-05}$ | 0.0012 |
| **East Africans meta-analysis** |  |  |  |
| Allele Freq. | 0.10 | 0.13 | 0.16 |
| Effect (Beta) | -0.32 | -0.17 | 0.25 |
| SE | 0.047 | 0.041 | 0.038 |
| P value | $3.04 \times 10^{-11}$ | $3.41 \times 10^{-05}$ | $5.06 \times 10^{-11}$ |
| **South Africans** |  |  |  |
| Allele Freq. | 0.066 | 0.19 | 0.200 |
| Effect (Beta) | -0.23 | -0.21 | 0.16 |
| SE | 0.11 | 0.073 | 0.072 |
| P value | 0.040 | 0.0037 | 0.029 |
| **DIASPORA AFRICAN ANCESTRY COHORTS** |  |  |  |
| **SCAALA (Brazil)** |  |  |  |
| Allele Freq. | 0.163 | 0.30 | 0.10 |
| Effect (Beta) | -0.075 | -0.12 | 0.17 |
| SE | 0.069 | 0.056 | 0.087 |
| P value | 0.28 | 0.039 | 0.050 |
| **JHS (USA)** |  |  |  |
| Allele Freq. | 0.10 | 0.15 | 0.17 |
| Effect (Beta) | -0.12 | -0.02 | 0.10 |
| SE | 0.038 | 0.031 | 0.030 |
| P value | 0.0016 | 0.56 | $3.97 \times 10^{-4}$ |
| **MESA (USA)** |  |  |  |
| Allele Freq. | 0.116 | 0.172 | 0.155 |
| Effect (Beta) | -0.218 | 0.0453 | 0.0188 |
| SE | 0.0610 | 0.0523 | 0.0530 |
| P value | 0.00036 | 0.39 | 0.72 |
| **All of Us (USA)** |  |  |  |
| Allele Freq. | 0.10 | 0.16 | 0.17 |
| Effect (Beta) | -0.124 | -0.0593 | 0.0717 |
| SE | 0.026 | 0.0218 | 0.0208 |
| P value | $2.18 \times 10^{-06}$ | 0.0065 | 0.00056 |
| **UK Biobank (UK)** |  |  |  |
| Allele Freq. | 0.09 | 0.15 | 0.16 |
| Effect (Beta) | -0.19 | -0.02 | 0.07 |
| SE | 0.03 | 0.02 | 0.02 |
| P value | $6.66 \times 10^{-12}$ | 0.48 | $6.40 \times 10^{-4}$ |
| **EUROPEAN ANCESTRY COHORT** |  |  |  |
| **UK Biobank (UK)** |  |  |  |
| Allele Freq. | 0.291 | 0.41 | n/a* |
| Effect (Beta) | -0.187 | -0.068 | n/a* |
| SE | 0.002 | 0.002 | n/a* |
| P value | $2.90 \times 10^{-1689}$ | $3.53 \times 10^{-266}$ | n/a* |

Abbreviation: ref. reference; SE standard error; freq. frequency; n/a not available. UKB European summary data was obtained from Minousaki et al<sup>3</sup>. The discovery association analyses were adjusted for age, sex and season, and incorporated GRM to account for relatedness. The rs7950991 variant was missing in European GWAS, it is rare in Europeans (MAF<0.001, dbSNP). However, rs12284909, which is in high linkage disequilibrium with rs7950991 in our discovery cohorts ( $r^2=0.98$ ), was genome-wide significant in Europeans (effect/other allele; A/G, A allele frequency=0.006, beta = -0.09, $P=7.6 \times 10^{-12}$ ).

**Supplementary Table 4.**
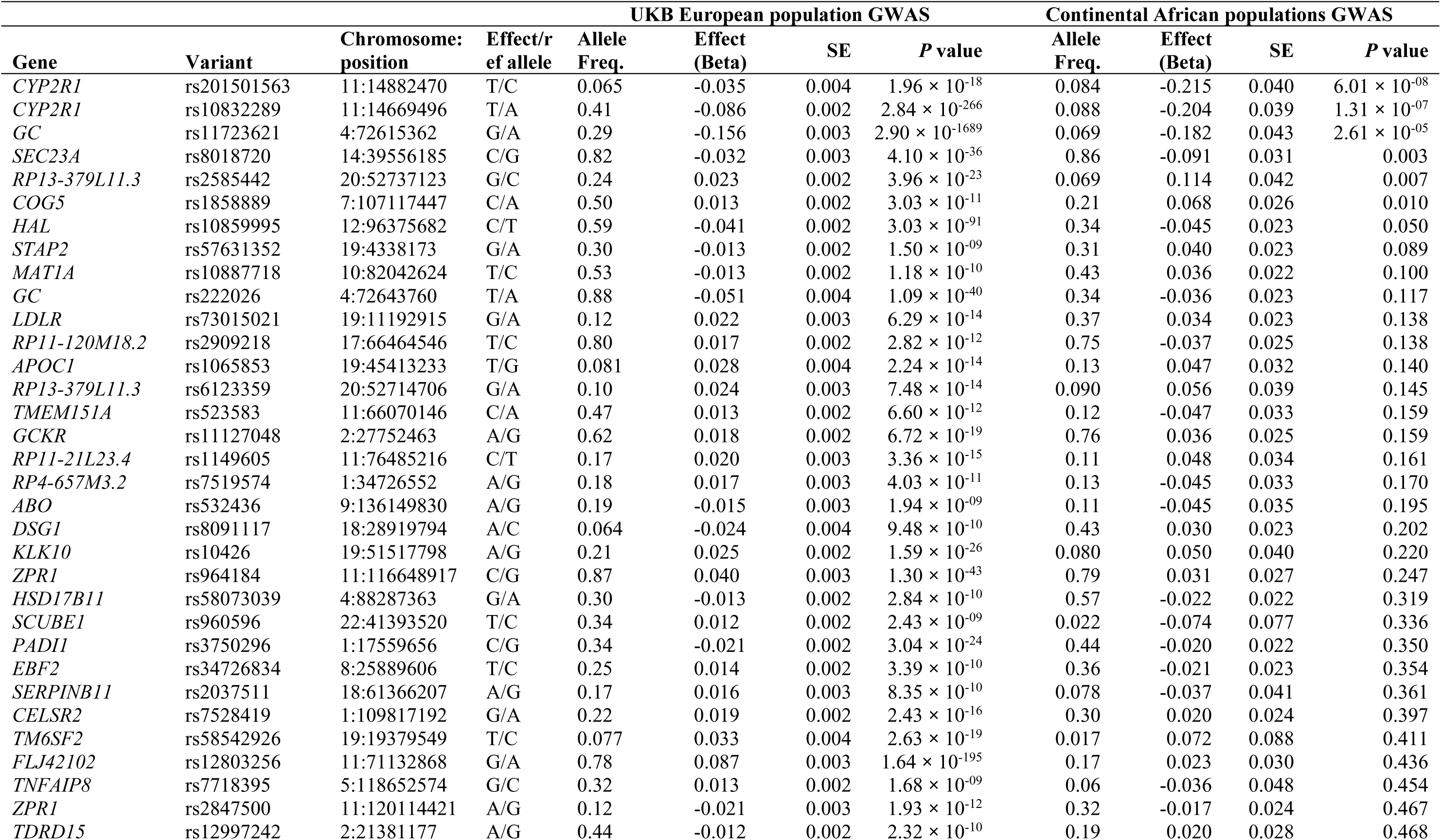
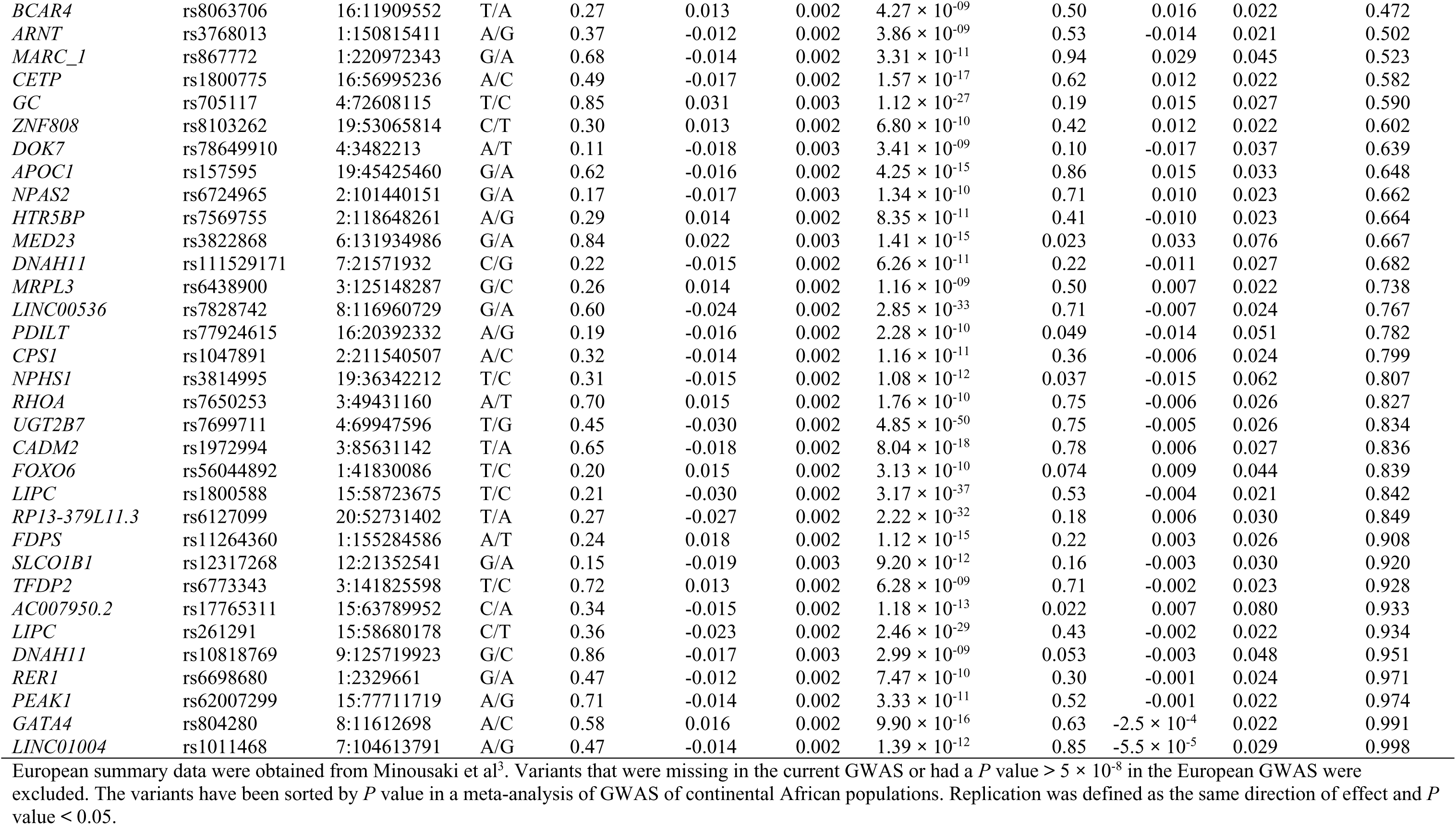
Replication of genome-wide significant variants in UK Biobank European-ancestry GWAS on 25(OH)D concentrations in a meta-analysis of continental African cohorts.

**Supplementary Table 5.** Effect of *OSBPL11* lead variant, rs2979356, on select biomarkers of lipid metabolism and cardiovascular health in African ancestry individuals.

|  | **n** | **Beta** | **SE** | **P** |
| --- | --- | --- | --- | --- |
| **HDL cholesterol** |  |  |  |  |
| AADM (overall) | 4,295 | 0.0338 | 0.0319 | 0.29 |
| AADM (West Africa) | 3,561 | -0.0272 | 0.0368 | 0.46 |
| AWI-Gen (overall) | 10,603 | 0.004 | 0.020 | 0.83 |
| AWI-Gen (West Africa) | 3,763 | -0.027 | 0.034 | 0.43 |
| UKB (African ancestry) | 5,754 | -0.00359 | 0.0240 | 0.055 |
| AMP-CMDKP | 128,439 | -0.0039 | 0.0039 | 0.47 |
| GLGC | 90,804 | 0.00209 | 0.00595 | 0.73 |
| **Total cholesterol** |  |  |  |  |
| AADM (overall) | 4,284 | 0.0154 | 0.0319 | 0.63 |
| AADM (West Africa) | 3,553 | 0.0188 | 0.0366 | 0.61 |
| AWI-Gen (overall) | 10,603 | -0.023 | 0.020 | 0.20 |
| AWI-Gen (West Africa) | 3,763 | -0.031 | 0.033 | 0.36 |
| UKB (African ancestry) | 6,212 | 0.0174 | 0.0248 | 0.32 |
| AMP-CMDKP | 128,439 | 0.0086 | 0.0039 | 0.11 |
| GLGC | 92,554 | -0.00928 | 0.00589 | 0.12 |
| **HOMA2-%S** (AADM) | 2,390 | -0.0140 | 0.047 | 0.77 |
| **HOMA2-B** (AADM) | 2,390 | -0.0181 | 0.036 | 0.61 |
| **TG/HDL ratio** (AWI-Gen West Africa) | 23,139 | 0.0277 | 0.0227 | 0.23 |
| **Fasting Glucose** (AWI-Gen West Africa) | 3,591 | 0.024 | 0.022 | 0.57 |
| **Fasting Insulin** (AWI-Gen West Africa) | 2,129 | 0.007 | 0.021 | 0.14 |

The effect/alternative allele is T/C. HOMA2-B estimates beta cell function and insulin production in the body using fasting insulin and glucose levels. HOMA2-%S is a calculation that measures how well peripheral tissues respond to insulin and absorb glucose from the bloodstream.\*The AWI-Gen Study (Africa Wits-INDEPTH Partnership for Genomic Studies) aimed to investigate the genetic and environmental factors influencing cardiometabolic diseases in sub-Saharan Africa, with participants recruited from Burkina Faso, Ghana, Kenya, Namibia, South Africa, and Tanzania<sup>37</sup>. AWI-Gen W. Africa included The Gambia and Ghana. The AMP-CMDKP (African American Cardiometabolic Disease Knowledge Portal) aimed to explore genetic contributors to cardiometabolic diseases, recruiting participants from diverse African ancestry populations (https://hugeamp.org/). Participants in the AADM study and the UK Biobank study are described in the methods.

**Supplementary Table 6.** Study cohort site, design, site, genotyping and phenotyping.

| Study name | Continental African discovery cohorts |  |  |  |  | African ancestry replication cohorts |  |  |  |  |
| --- | --- | --- | --- | --- | --- | --- | --- | --- | --- | --- |
|  | Kilifi immunology cohort (Kenya) | The Entebbe Mother and Baby Study (Uganda) | The VAC050 ME-TRAP malaria vaccine trial (Burkina Faso) | Malaria study (The Gambia) | Soweto Vaccine Response Study (South Africa) | Jackson Heart Study (USA) | Multi-Ethnic Study of Atherosclerosis (USA) | UK Biobank African ancestry participants (UK) | SCAALA (Brazil) | All of US Research Program (USA) |
| Study Site | Kilifi, Kenya | Entebbe, Uganda | Banfora, Burkina Faso | West Kiang, The Gambia | Soweto, South Africa | Jackson, Mississippi, USA | USA | UK | Salvador, Brazil | USA |
| Study design | Community-based cohort | Community-based cohort | Community-based cohort | Community-based cohort | Community-based cohort | Community-based cohort | Community-based cohort | Population cohort | Community-based cohort | Population cohort |
| Genotyping platform | H3Africa chip v1 | Illumina HumanOmni 2.5M-8 | Illumina HumanOmni 2.5M-8 | H3Africa chip v1 | Illumina HumanOmni 2.5M-8 | Affymetrix 6.0 | Affymetrix 6.0 | UK Biobank Axiom™ Array | Illumina HumanOmni 2.5M | Illumina NovaSeq 6000 whole genome sequencing |
| Number of variants in chip | 2.5 million | 2.5 million | 2.5 million | 2.5 million | 2.5 million | 906,600 | 906,600 | 850,000 | 2.5 million | Whole genome sequence |
| Vitamin D assay | CMIA (Abbott Architect, USA) | CMIA (Abbott Architect, USA) | CMIA (Abbott Architect, USA) | CMIA (Abbott Architect, USA) | CMIA (Abbott Architect, USA) | LC-MS-MS | LC-MS-MS | DiaSorin Liaison XL assay | Inhibitory enzyme immunosorbent assay (Bolton, UK) | Multiple assays |
| CMIA, chemiluminescent microparticle immunoassay; SCAALA, Social Change Asthma and Allergy; UKB, UK Biobank; LC-MS-MS, Liquid chromatography with tandem mass spectrometry. |  |  |  |  |  |  |  |  |  |  |

**Supplementary Figure 1.**
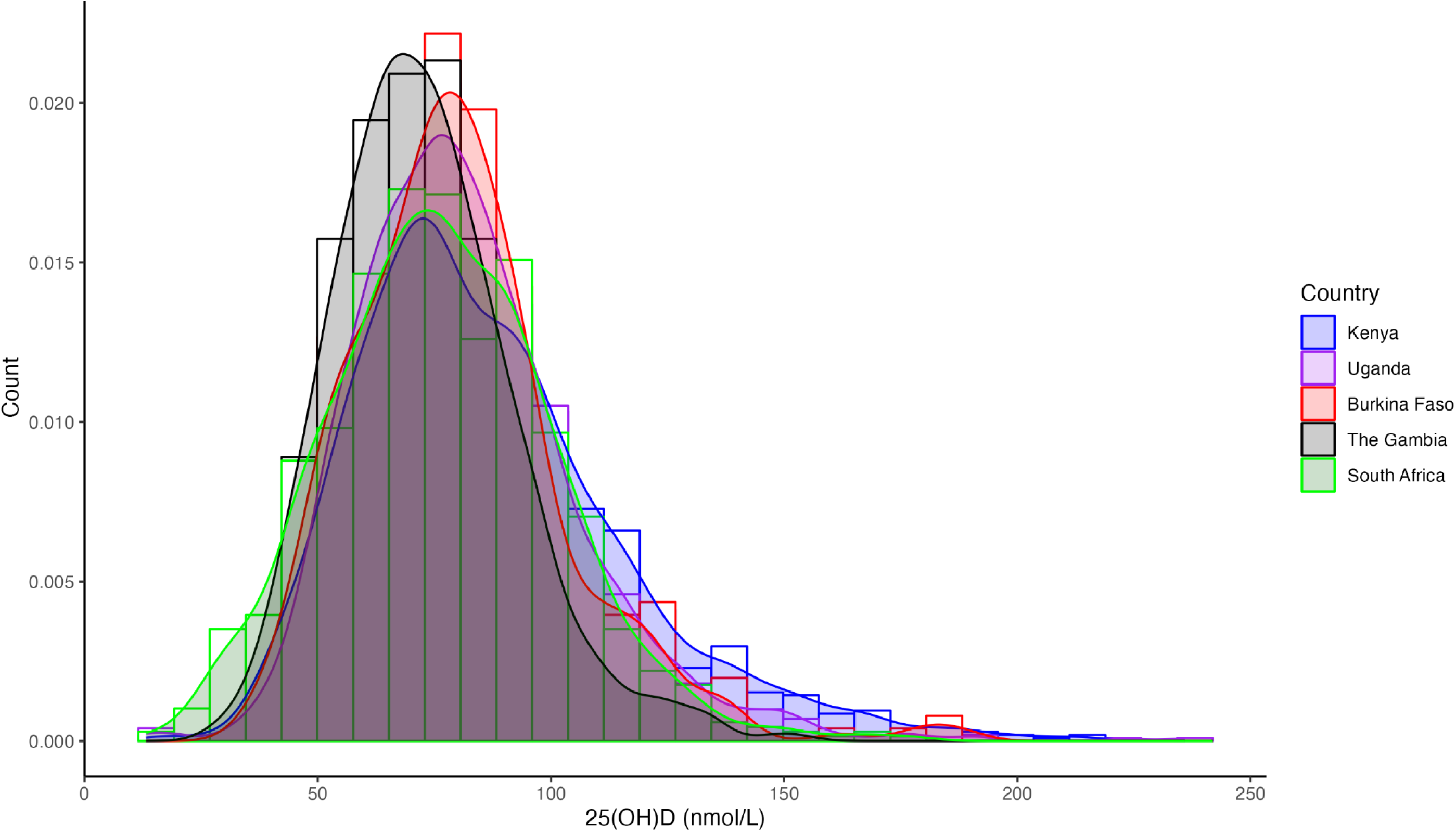
Histogram with density plots showing the distribution of 25(OH)D concentrations. Shapiro-Wilk’s test for normality indicated that 25(OH)D concentrations were skewed (*W*=0.95, *P* value < 0.001).

**Supplementary Figure 2.**
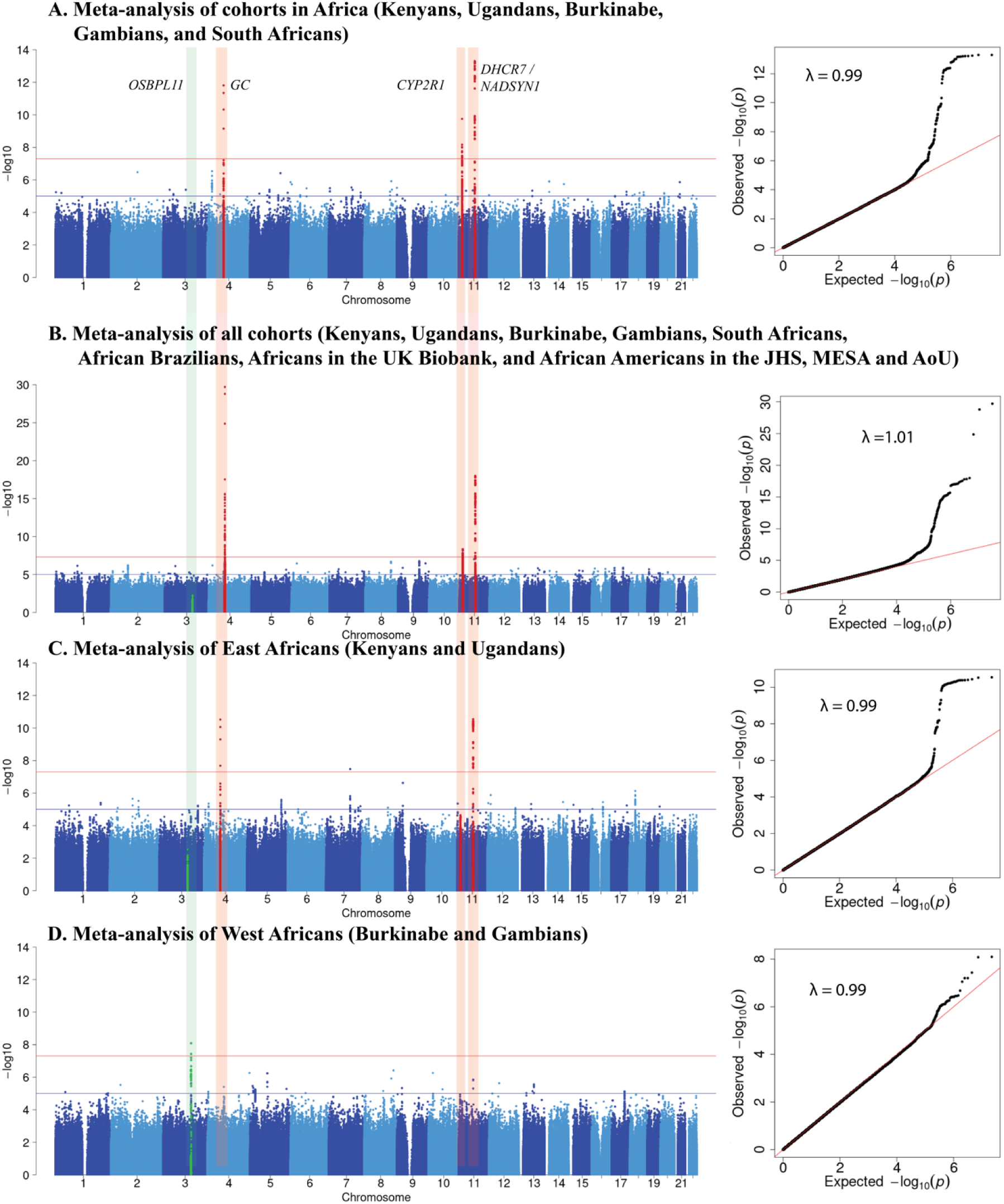
Manhattan and QQ plots of GWAS of 25(OH)D concentrations meta-analysis of all cohorts in Africa (A), all discovery and replication cohorts (B), East African cohorts (C), and West African cohorts (D). Variants in known loci (*GC, CYP2R1/PDE3B* and *DHCR7/NADSYN1*) are indicated in red and novel loci (*OSBPL11*) in green.

**Supplementary Figure 3.**
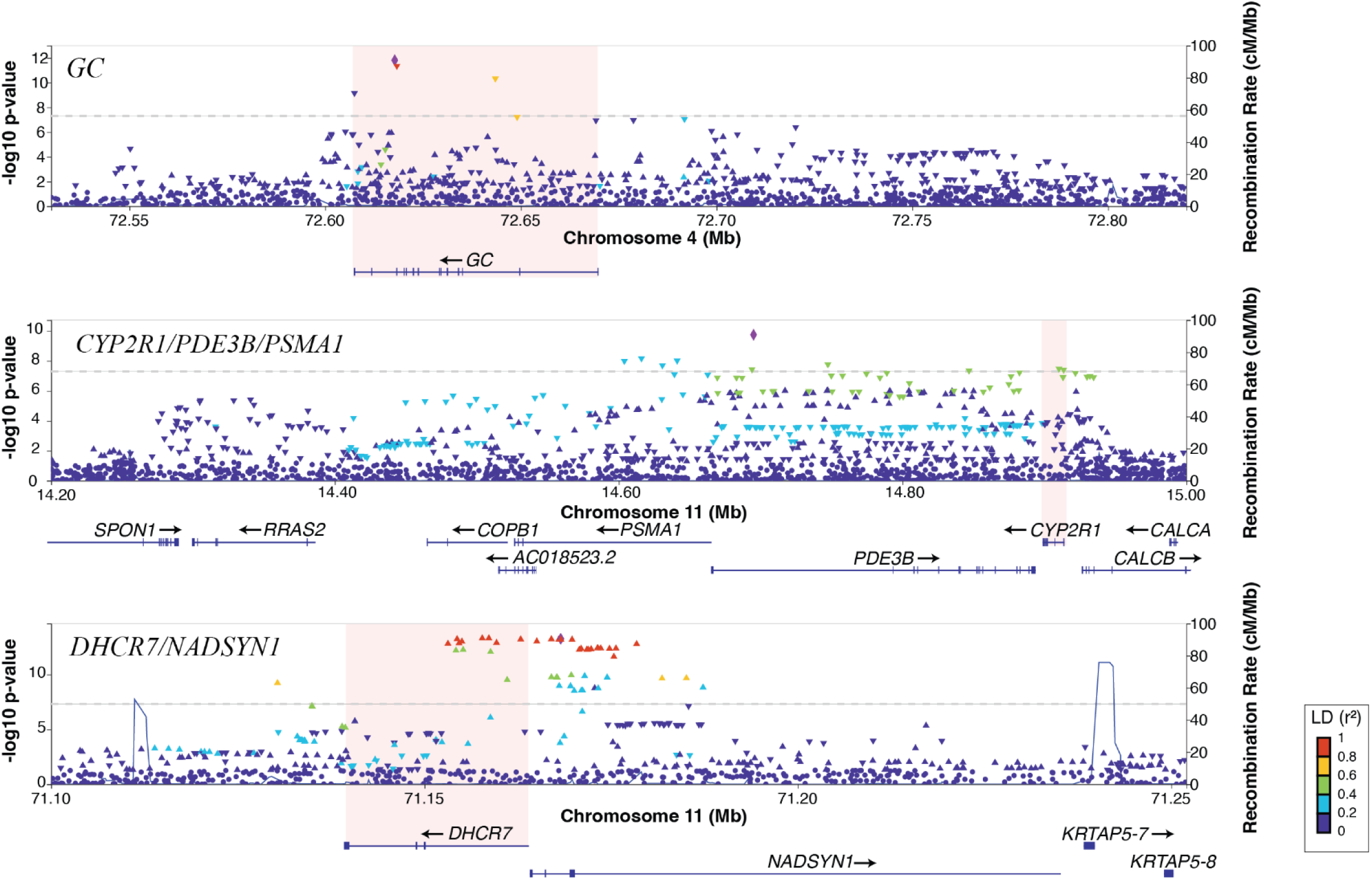
Regional association plots for the *GC, CYP2R1/PDE3B/PSMA1* and *DHCR7*/*NADSYN1* loci. The top variant in the regional plots is shown with a purple diamond shape and the rest are coloured according to their LD (R^2^) relative to the top variant in individuals of African ancestry in this study; colour varies between absent LD (blue) and total LD (red). The direction of the triangle of the top variant in each locus in the regional plots indicates the direction of effect on 25(OH)D concentrations.

**Supplementary Figure 4.**
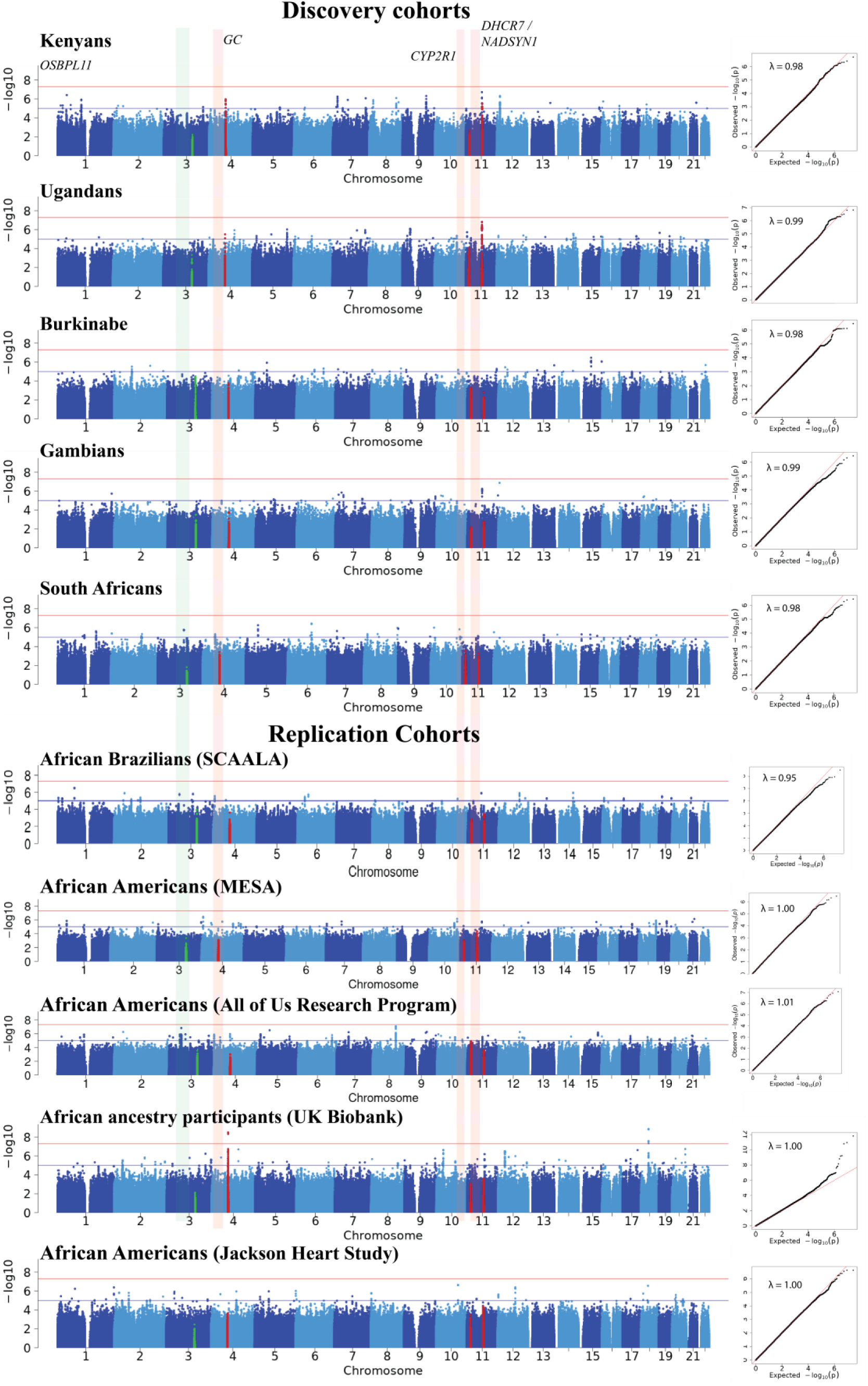
Manhattan and QQ plots of GWAS of 25(OH)D concentrations by cohort. Variants in and close to known loci (*GC, CYP2R1/PDE3B* and *DHCR7/NADSYN1*) are shown in red and novel loci (*OSBPL11*) are shown in green.

**Supplementary Figure 5.**
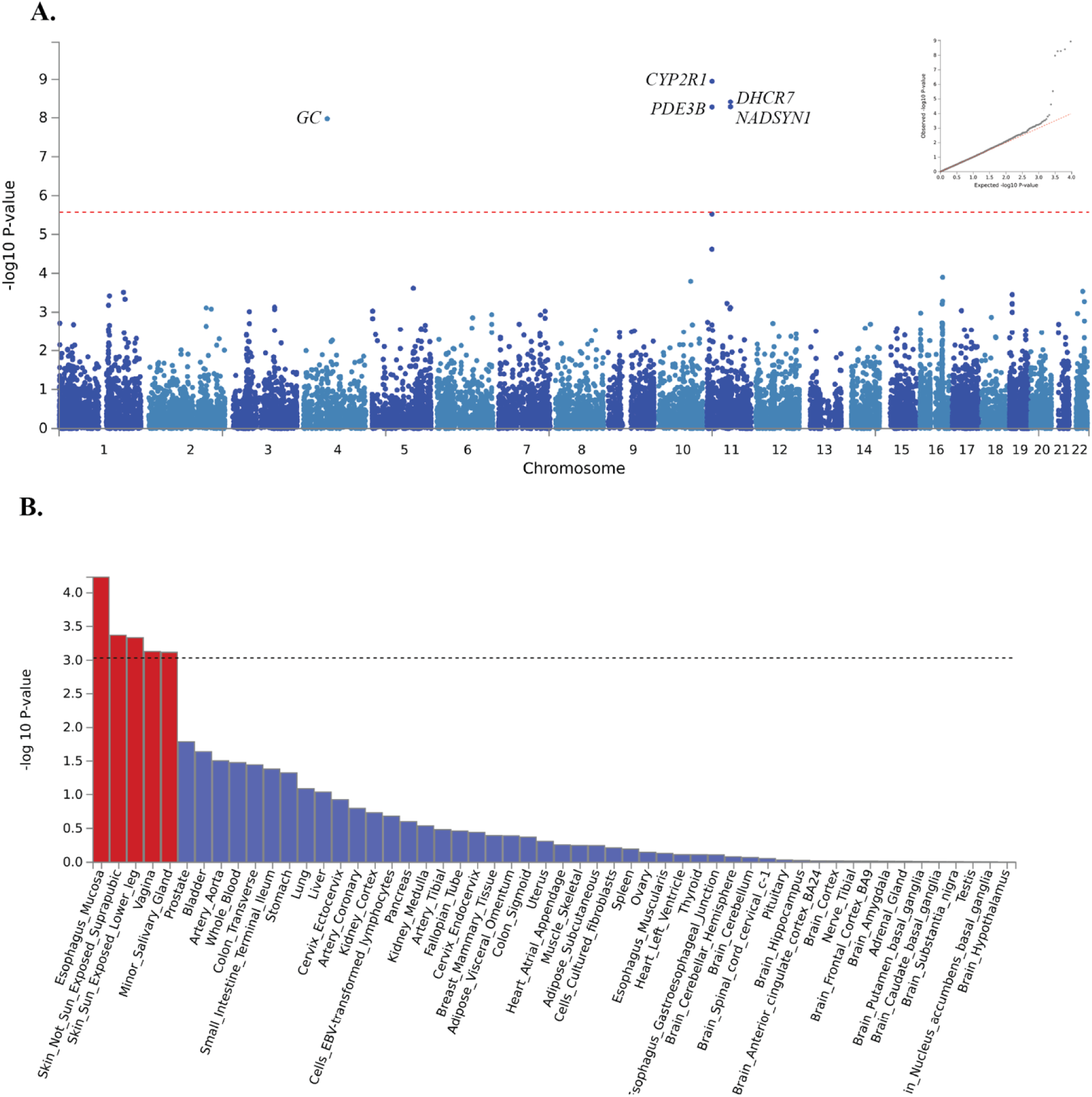
Manhattan plot of MAGMA gene-based association test results and corresponding QQ plot (A) and MAGMA tissue expression analysis, where tissues that exceed the significance threshold (P = 0.05/18912 = 2.64 × 10^-6^) are shown in red (B), Dark blue areas highlight genomic risk loci. Green lines represent eQTL associations, while orange lines depict chromatin interactions. Genes mapped by both eQTL and chromatin interactions are highlighted in red. The gene mapping was conducted using FUMA using African ancestry data (https://fuma.ctglab.nl/) with the gene-based test performed via MAGMA (v1.08) ^39^. Input variants were mapped to 18,912 protein-coding genes.

**Supplementary Figure 6.**
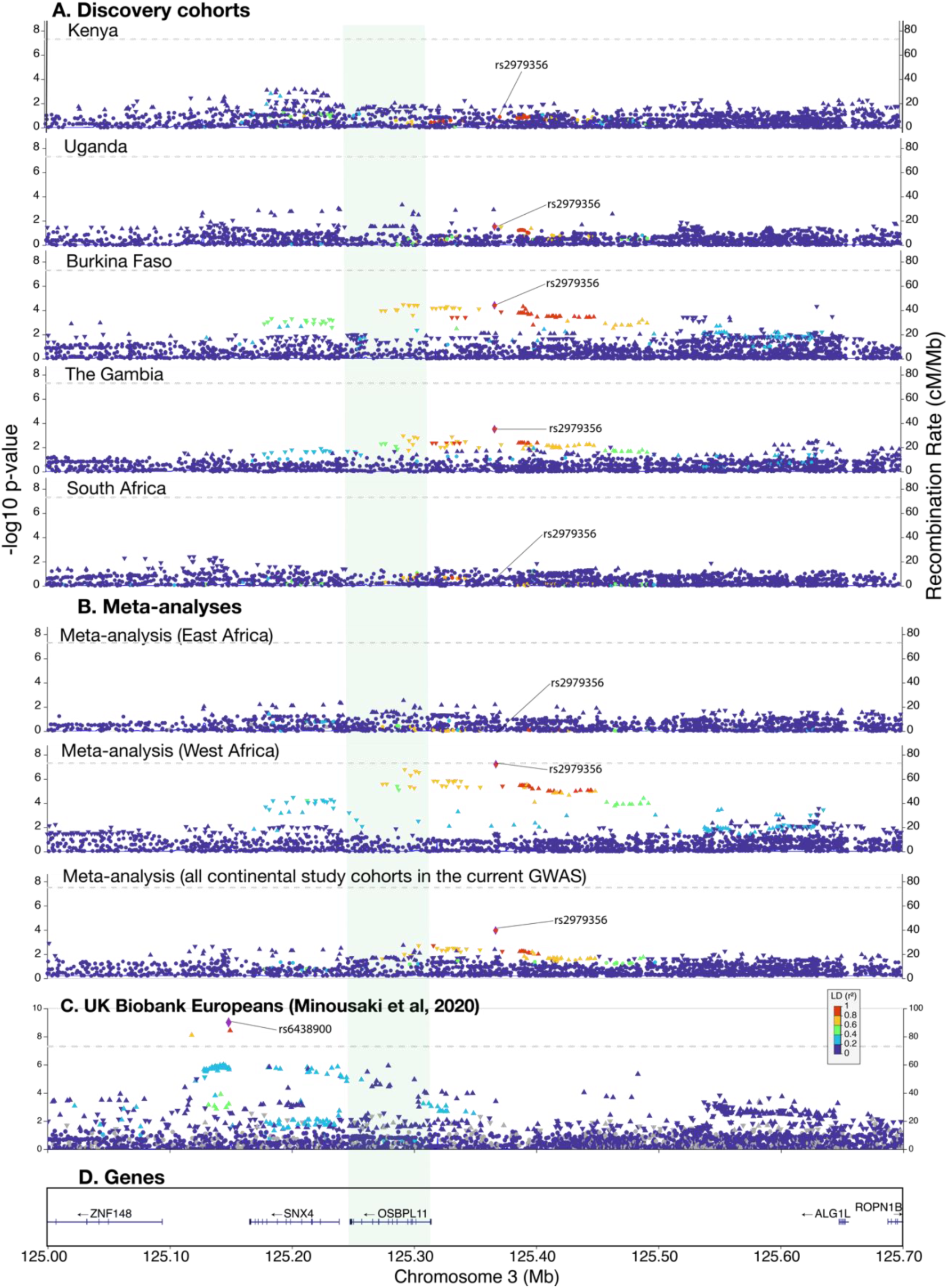
Regional association plots for the *OSBPL11* locus in chromosome 3 for individual continental African cohorts (A), meta-analyses (B), UK Biobank Europeans (C), and genes in the regions (D). The direction of the triangle of the top variant indicates the direction of effect on 25(OH)D concentrations. The variants are coloured according to their LD (R^2^) relative to the top variant. The reference variant (rs2979356) is illustrated in a purple diamond shape.

**Supplementary Figure 7.**
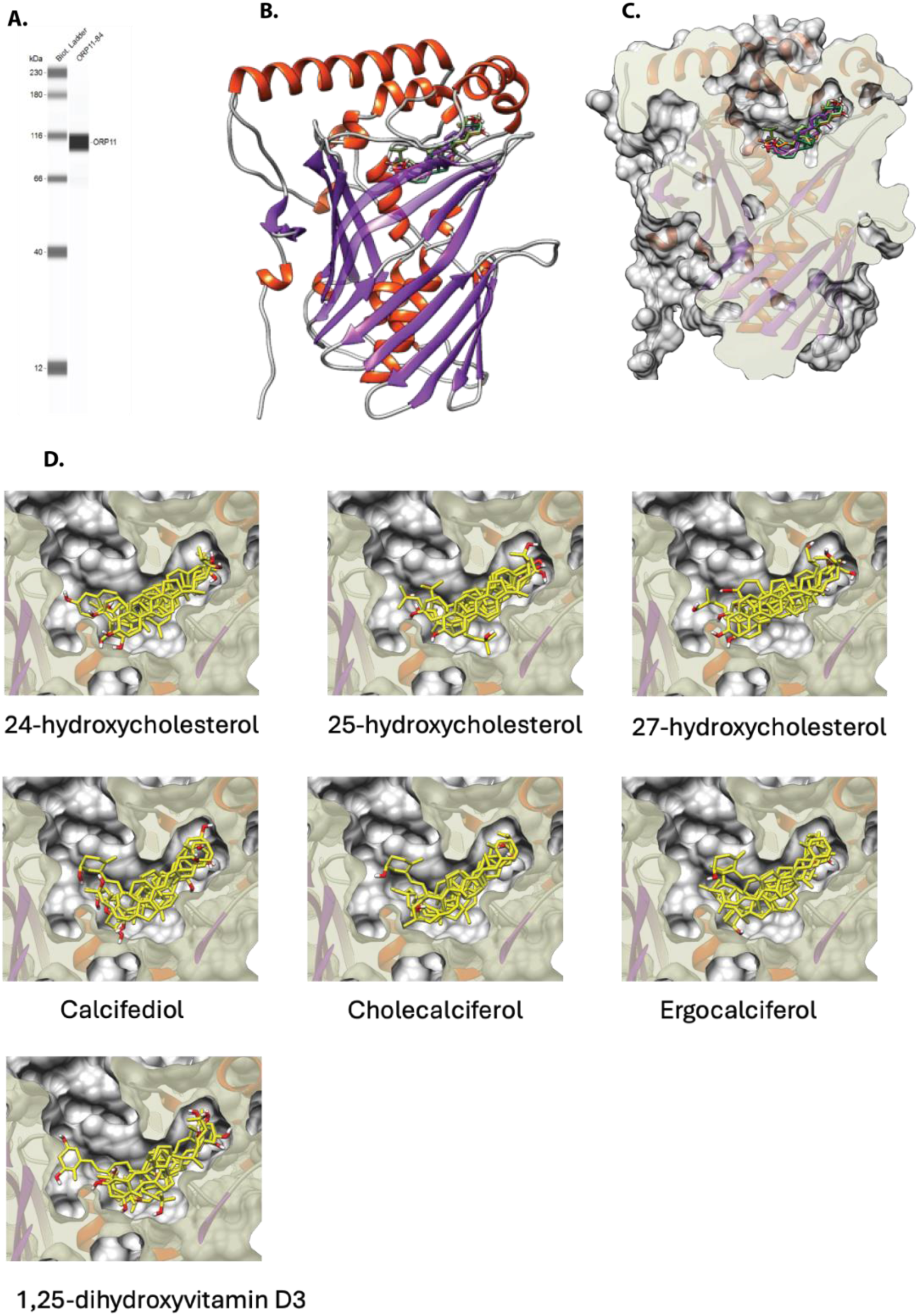
OSBPL11 Alpha Fold model. Protein immunoassay of OSBPL11-myc-His overexpressed 293T lysate (A), OSBPL11 AlphaFold model (B), molecular docking of oxysterols and vitamin D compounds into oxysterol binding site in the AlphaFold modeled OSBPL11 (C), visualization of all binding poses for individual sterols and vitamin D analogs (D). Docking was performed using AutoDock Vina (v. 4.2.6, see Web Resources) plugin through UCSF Chimera (v. 1.17.3). Figures were made in UCSF Chimera (see Web Resources).

**Supplementary Figure 8.**
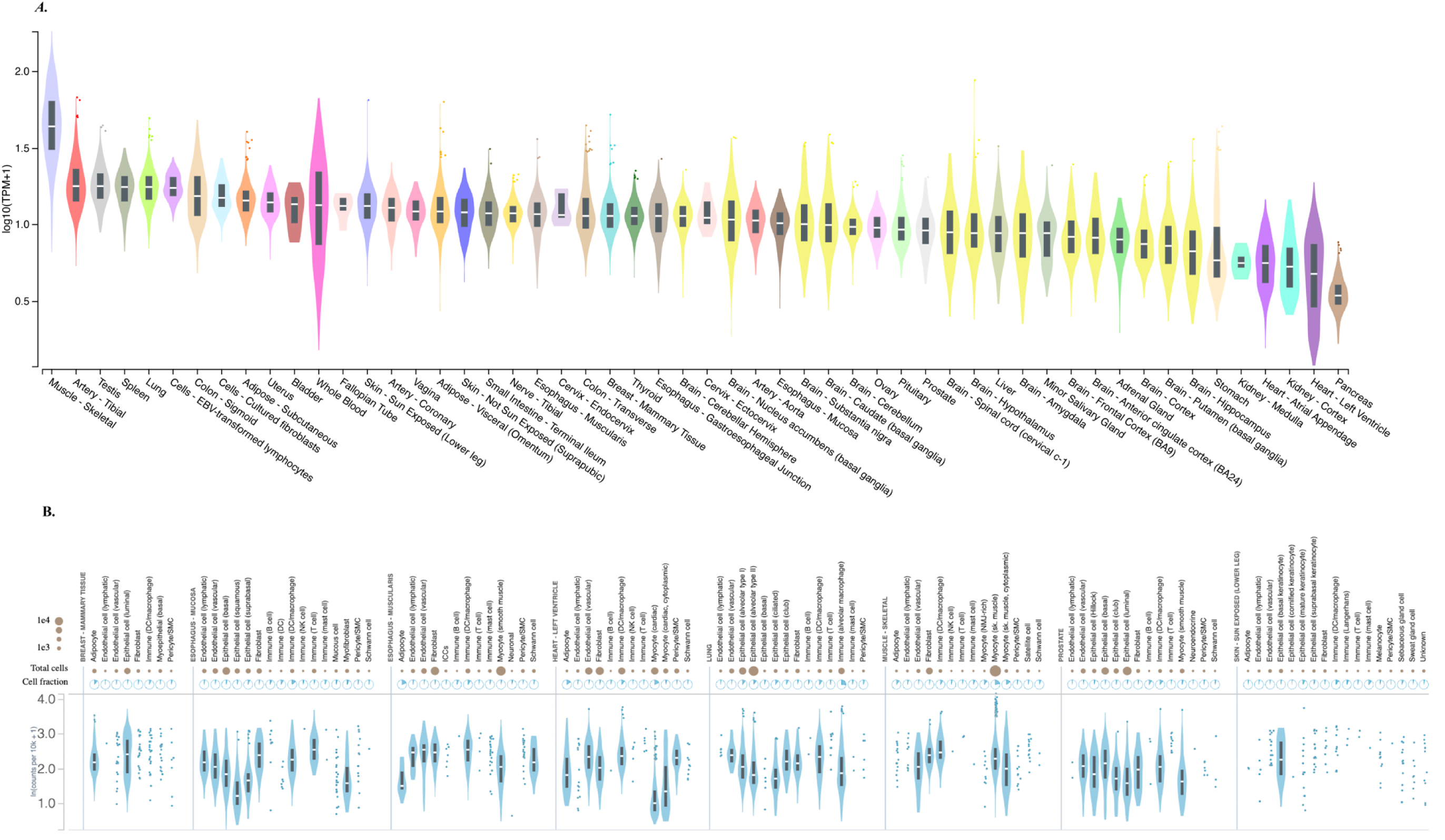
Bulk tissue (A) and single-cell (B) gene expression for *OSBPL11* (ENSG00000144909.7) Gene expression data were obtained from GTEX Portal version V8 (https://gtexportal.org/).

**Supplementary Figure 9.**
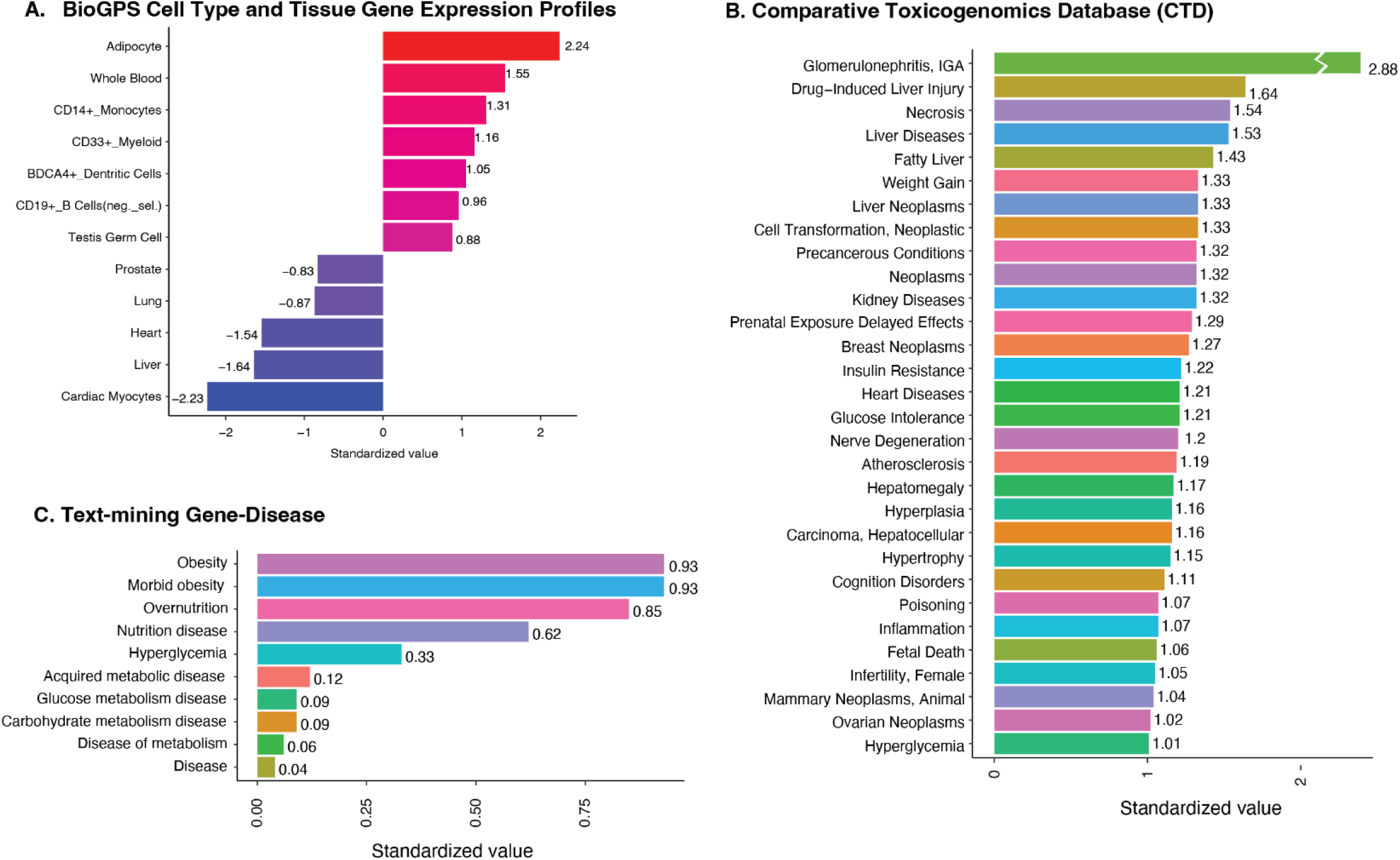
Select *OSBPL11* search results from Harmonizome portal: differential gene-expression profiles in human tissues/cells (A) and mouse cell types in BioGPS datasets (B), GEO Signatures of Differentially Expressed Genes for Diseases (C), association of *OSBPL11* with diseases in Comparative Toxicogenomics Database (CTD) (D) and Text-mining Gene-Disease dataset (E). Complete Harmonizome (https://maayanlab.cloud/Harmonizome/) search results are presented in Supplementary Dataset 6. The standardized values shown represent z-scores calculated in the Harmonizome database, indicating how many standard deviations each gene’s measurement is from the mean of the dataset, thus allowing for direct comparison across different datasets.

**Supplementary Figure 10.**
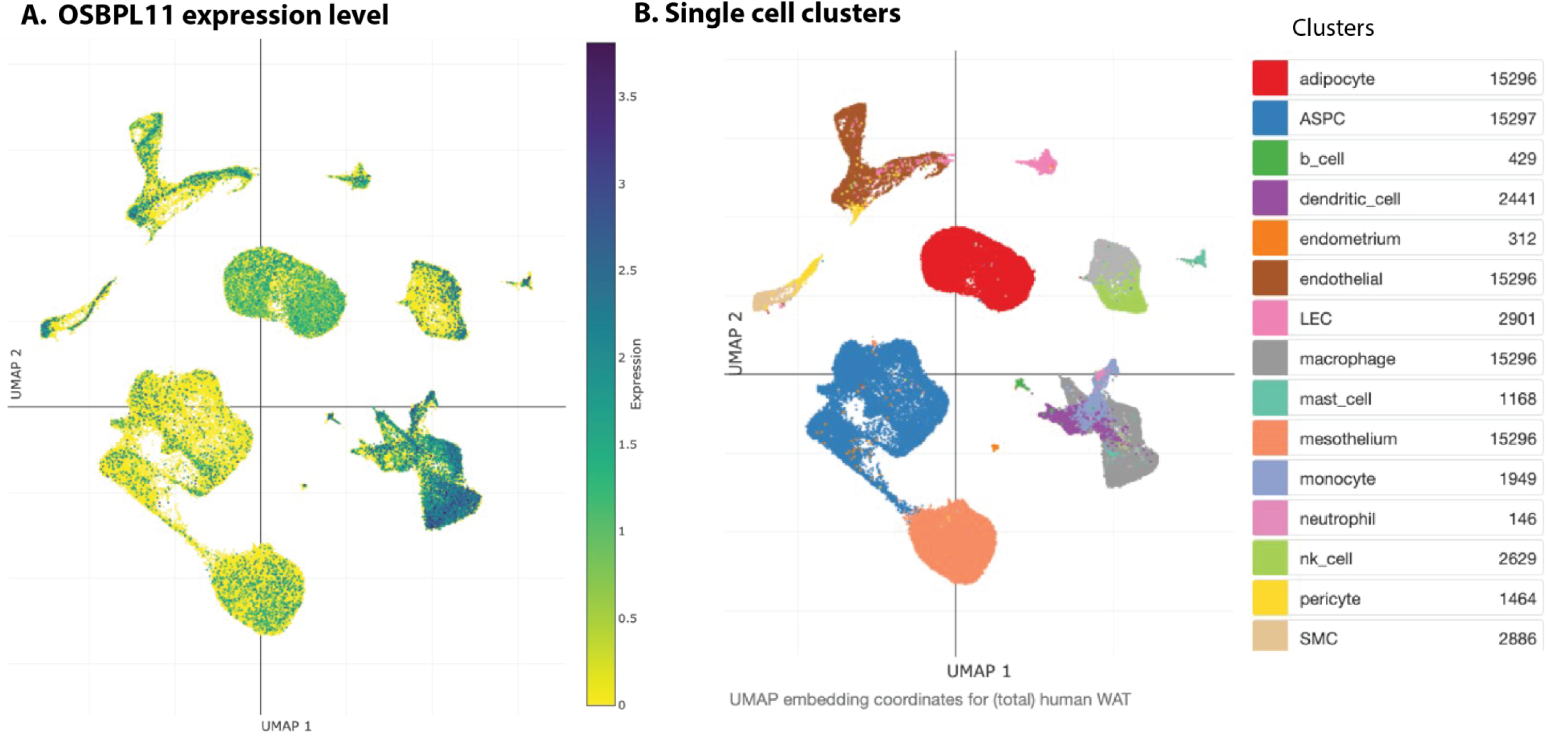
Single cell atlas of human white adipose tissue showing expression levels for *OSBPL11*. Data was retrieved from https://singlecell.broadinstitute.org/single_cell/study/SCP1376/a-single-cell-atlas-of-human-and-mouse-white-adipose-tissue. Abbreviations: OSBPL11, oxysterol binding protein like 11; UMAP, Uniform Manifold Approximation and Projection; B cell, B lymphocyte; T cell, T lymphocyte; LEC, lymphatic endothelial cell; NK cell, natural killer cell; SMC, smooth muscle cell; ASPC, adipose stem and progenitor cell.

**Supplementary Figure 11.**
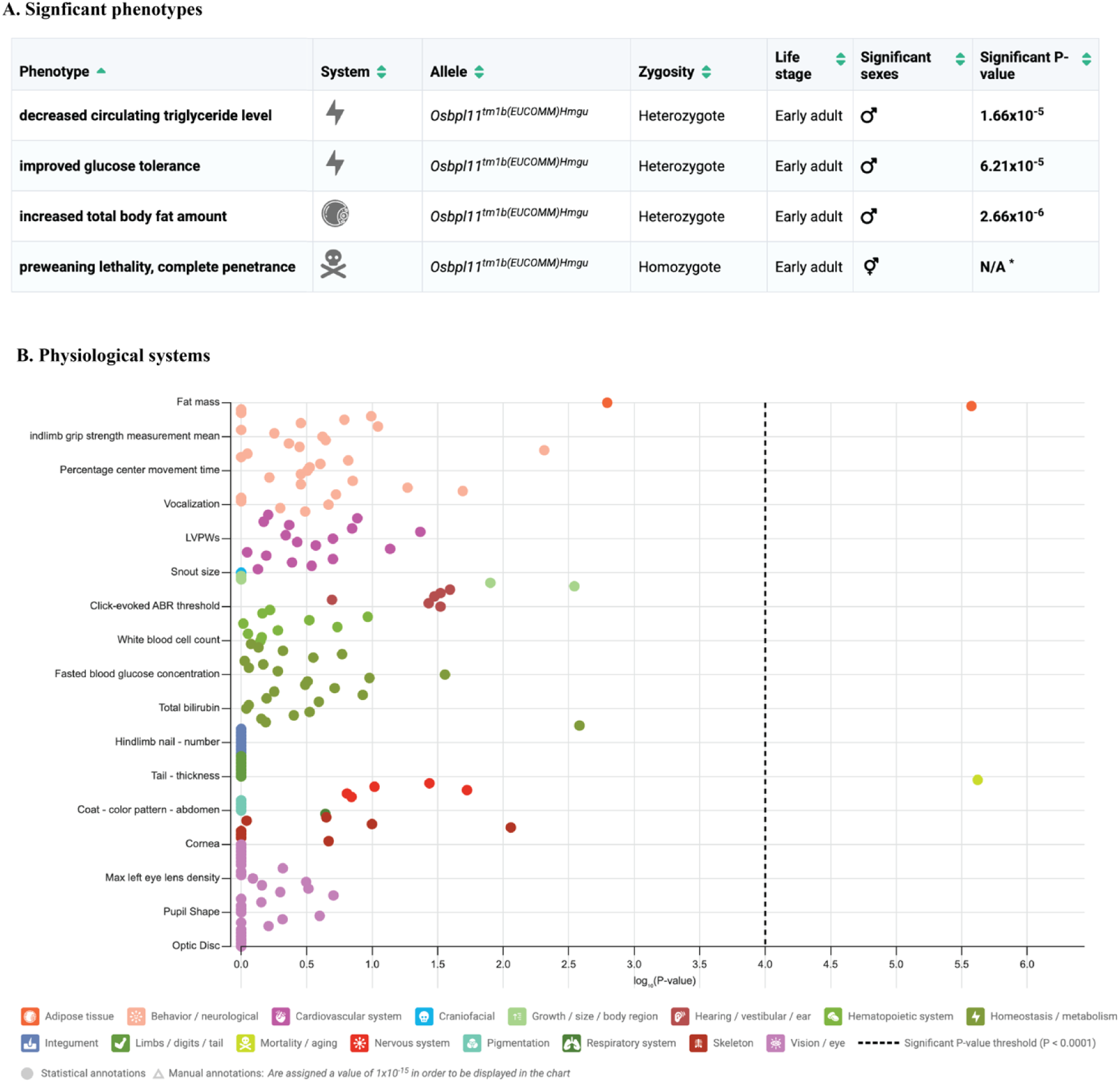
Phenotypes with significant changes (A) and phenotyping screening results of Osbpl11 mouse knockout mice (B). The International Mouse Phenotyping Consortium (IMPC) applies a panel of phenotyping screens to characterize single-gene knockout mice in comparison to wild types. Data sourced from the IMPC (https://www.mousephenotype.org/data/genes/MGI:2146553).

## Methods

### Ethics statement

Written informed consent was obtained from the parents or guardians of each child prior to their enrolment in this study. Study protocols received approval from the local institutional review boards at each participating site.

### Study participants (Discovery GWAS)

This GWAS study included young community-based children living in Kenya (n = 1361), Uganda (n = 1301), Burkina Faso (n = 329), The Gambia (n = 629), and South Africa (n = 889), as previously described^6^. Details of these cohorts are briefly described below.

*Kilifi, Kenya (3.5° S, 39.9° E)* –This is an ongoing community-based cohort aimed at evaluating immunity to malaria in children from birth to 8 years^40^. The children were followed up weekly for malaria and other infections, and anthropometric measurements and blood samples were collected during annual cross-sectional surveys.

*Entebbe, Uganda (0.1° N, 32.5° E)* –This cohort was part of the Entebbe Mother and Baby Study (EMaBS) which was a prospective birth cohort study that was originally designed as a randomized controlled trial (ISRCTN32849447) to evaluate the effects of helminths and anthelminthic treatment during pregnancy and early childhood on immunological responses to routine vaccinations. Blood samples and anthropometric measurements were collected at birth and at subsequent annual visits up to age five years^41^.

*Banfora, Burkina Faso (10.6° N, 4.8° W)* –This cohort participated in the VAC050 ME-TRAP Malaria Vaccine trial aimed at testing the effectiveness, safety, and immunogenicity of a prime-boost liver-stage malaria vaccine in children aged between six and 17 months^42^. Blood samples were collected, and anthropometric measurements taken at multiple timepoints after receipt of the experimental vaccine based on another study investigating the genetics of responses against childhood vaccines^41^.

*West Kiang, The Gambia (13.3° N, 16.0° W)* –This was a community-based cohort study aimed at investigating the effect of genetic variants on anaemia during the malaria season in children aged between two and six years from 10 rural Gambian villages ^43^. Blood samples and anthropometric measurements from the start of the malaria season were included in this study.

*Soweto, South Africa (26.2° S, 27.9° E)*

The Soweto Vaccine Response Study recruited infants from vaccine trials coordinated by the Respiratory and Meningeal Pathogens Unit (http://www.rmpru.com/). The study used stored plasma samples collected at 12 months of age from infants that had received all their routine vaccines. Soweto, in South Africa is a non-malaria endemic region and anthropometry was not measured in this cohort^41^.

### Genotyping and imputation

The Ugandan (n=1390), Burkinabe (n=354) and South African (n=749) discovery cohorts were genotyped on the Illumina HumanOmni 2.5 M-8 (“octo”) BeadChip array version 1.1 as part of GWAS (Illumina Inc., San Diego, USA), while the Kenyan (n=1241) and Gambian cohorts (n=550) were genotyped on the H3A Custom Genotyping Array v1.0 (H3Africa v1.0 contains approximately 2.5 million variants that are optimized for African ancestry populations). Variant quality control (QC) was performed separately for each cohort using the H3Africa GWAS pipeline version 3 (see Web Resources) to exclude samples with genotype call rate < 97%, heterozygosity >3 SDs around the mean, discrepancy between genotype and allocated sex, and related individuals (identity by descent >0.9). Thereafter, variants passing the following filters were retained: >97% call rate, minor allele frequency >0.01 and Hardy–Weinberg equilibrium (HWE) >0.008. Imputation was performed using the African Genome Resources (AGR) Haplotype Reference Panel using the Sanger Imputation Service (see Web Resources). The AGR haplotype reference panel is based on 4956 samples from African and non-African 1000 Genomes Phase 3 populations, and 2298 additional African genomes from Uganda, Ethiopia, Egypt, Namibia, and South Africa, resulting in an increased number of imputed variants, improved information score and imputation accuracy compared to the 1000 Genomes Phase 3 Version 5 or TOPMED imputation panels. We filtered the resulting imputation dataset for variants with information score (info) ≥ 0.3 and MAF ≥ 0.01 for association analyses.

### Laboratory assays

25(OH)D was measured using a chemiluminescent microparticle immunoassay (CMIA, Abbot Architect) in the discovery cohorts (Supplementary Table 6). The CMIA 25(OH)D assay has comparable results to the liquid chromatography tandem mass spectrometry LC-MS/MS method^44^. Overall assay performance ranged from 2.8% to 7.9% for mean 25(OH)D concentrations ranging from 21 to 116 nmol/L. External quality assurance (DEQAS) data showed the 25(OH)D assay to have a mean (SD) bias of −2.7% (7.6) against the all-laboratory trended values, and one of −0.4% (7.7) against the target values. C-reactive protein (CRP) was measured using the MULTIGENT CRP Vario assay (Abbot Architect) and α1-antichymotrypsin (ACT) using immunoturbidimetry (Cobas Mira Plus Bioanalyser, Roche) in plasma samples stored at -80 °C (Supplementary Table 1). Malaria parasitaemia was detected using Giemsa-stained blood smears.

### Genome-wide association study (GWAS) analysis

To identify variants associated with 25(OH)D concentrations, we performed a linear mixed model GWAS within each study cohort as implemented in Genome-wide Complex Trait Analysis software version 1.24.4 (GCTA)^45^. GCTA uses a sparse genomic relationship matrix (GRM) to account for genetic structure within the cohort. The GRM was generated from unimputed genotype data using GCTA. To normalise 25(OH)D concentrations we rank-based inverse-normal transformed (RINT) 25(OH)D concentrations in the model and included age, sex, and season (at the time of vitamin D measurement) as covariates. We adjusted for season since seasonality is associated with vitamin D status in children living in Africa^6^. BMI was not included as it was not associated with vitamin D status in this population^6^. We computed and plotted PCs using PLINK software (version 2.0) to identify and visualize population structure and identify potential outliers and admixed individuals within the discovery cohorts. Vitamin D supplementation is rare in children living in rural African communities, and these data were not available. To identify conditionally independent associations at each genome-wide significant locus, we performed conditional and joint analyses using GCTA-COJO version 1.24.4^46^ using our GWAS summary data. We performed the association analyses using GCTA “cojo-slct” option conditioning on the lead variant and repeated the process if there was a secondary signal that reached genome-wide significance. We considered variants that reached genome-wide significance before and after conditioning to be conditionally independent. We used an inverse-variance-weighted fixed effect approach as implemented in METASOFT ^47^ (see Web Resources) to meta-analyse effect estimates across the study cohorts.

### Statistical power estimates

The power of the study for discovery was estimated using *Quanto*^48^. For a variant with a minor allele frequency (MAF) of 0.05, the study had 80% power to detect a genetic effect size (GES) of 8.6 (r^2^ of 0.011) and 97.7% power to detect a GES of 10.0 (r^2^ of 0.015). For a variant with a MAF of 0.10, the study has 80% power to detect a GES of 6.2 and 95% power to detect a GES of 7.0.

### Replication in African ancestry cohorts

To evaluate replication of our findings we performed a linear mixed model GWAS as implemented in GCTA in community-based cohorts of participants of African ancestry from the Social Change Asthma and Allergy in Latin America study (SCAALA), the Jackson Heart Study (JHS), the UK Biobank and the All of Us Research Program (Supplementary Table 1). The SCAALA Cohort (Salvador, Bahia, Brazil (12.9° S, 38.5° W)) is a longitudinal study in children initiated in 1997 aimed at assessing the impact of sanitation on health^49^. By 2013, the cohort included 1,206 participants aged 11-19 years, with 750 individuals selected for this analysis. The JHS is an ongoing community-based observational study with 5306 African American adult participants (Jackson, USA, 32.3° N, 90.2° W) aimed at identifying the environmental and genetic risk factors for the development of cardiovascular diseases among an African American population (see Web Resources). Details of the design, recruitment and initial characterization of the JHS study has been described in detail elsewhere^50^. Concentrations of 25(OH)D and CRP were measured in samples collected from clinic visits from 2000-2004. The Multi-Ethnic Study of Atherosclerosis (MESA) is a community-based cohort study initiated in 2000 to investigate subclinical cardiovascular disease. It includes 6,814 participants aged 45-84 years from six US field centers with African American, Hispanic, Chinese, and White ancestry, all free of cardiovascular disease at baseline. African ancestry participants were included in this analysis for replication (dbGaP accession phs000209.v13.p3). The UK Biobank (UKB, n=500,000) is a prospective cohort study following the health and well-being of middle-aged and older adults recruited throughout the UK between 2006 and 2010^51^. Concentrations of 25(OH)D and CRP and anthropometry were measured during a baseline evaluation at UKB assessment centres. We performed imputation, quality control, and association analyses using the same parameters and software as for the discovery GWAS. We performed a linear mixed model GWAS (with GRM) as implemented in GCTA for the replication cohorts and adjusted for cohort-specific covariates linked to vitamin D levels: age and sex for SCAALA and age, sex, season, vitamin D supplementation and BMI for UKB and JHS (assessment centre also included for the UKB). Additionally, we conducted a meta-analysis including both the discovery and replication cohorts (Brazilians in SCAALA, African American participants in the JHS and African ancestry participants in the UKB). The All of Us Research Program is a nationwide initiative in the United States aiming to collect health data from one million or more participants to accelerate research and improve health outcomes^52^. This program places a strong emphasis on diversity, with over 80% of participants coming from groups historically underrepresented in biomedical research, including significant representation of individuals of African ancestry. Since the GWAS for the All of Us study had already been performed, we incorporated the summary statistics obtained from their ALL by ALL platform into our analysis^52^. Participants in JHS and MESA were selected based on self-reported race, without exclusion of ancestry outliers. While self-reported population descriptors reflect social identity and should not be interpreted as proxies for genetic ancestry, prior studies have shown that most participants in both cohorts exhibit high proportions of African ancestry^53^. Analyses of the All of Us Research Program and UK Biobank have used genetically inferred African ancestry to characterize participants.

### Fine-mapping and functional annotations

We used the FUMA online pipeline version 1.4.1^54^ to annotate the GWAS results. We defined independent significant variants as variants with *P* < 5 × 10^-8^ and a pairwise r^2^ ≤ 0.6. We further defined lead variants from the independent significant variants as variants with r^2^ < 0.1. Genomic risk loci were defined by clustering independent significant SNPs that were in LD (r^2^ ≥ 0.1) and <250 kb apart. Each locus is represented by the top lead variant which has the minimum *P*-value in the locus. Genetic data from African populations in the 1000G phase3 were used as reference data to conduct LD analyses in FUMA. Genomic loci were mapped to genes using positional mapping (loci located in the genes or < 10 kb away) and expression quantitative trait locus (eQTL) mapping (loci were mapped to a gene if the corresponding variants had a significant effect on the expression of the gene using a false discovery rate (FDR) of < 0.05). eQTL mapping was performed using data from 27 tissues (Genotype-Tissue Expression (GTEx) version 8 Whole Blood and Minor Salivary Gland^55^, Database of Immune Cell Expression (DICE) ^56^, single-cell RNA eQTL ^57^ and Biobank-Based Integrative Omics Study (BIOS) QTL Browser^58^). Additionally, we searched for lead variants (rs2979356) and *OSBPL11* gene expression profiles in 54 human tissues and organs in the GTEx portal v8 ^59^ (https://gtexportal.org/). In addition, we also retrieved the expression profile of *OSBPL11* in the single cell atlas of human and mouse white adipose (see Web Resources).

To identify the most likely causal variant(s) for the four independent loci, we subsequently utilized FINEMAP to estimate the number of most likely causal variants per locus and generated a 95% credible set for each causal variant^60^. We used PolyPhen, a tool designed for predicting the possible impact of amino acid substitutions on the structure and function of human proteins, to systematically examine the credible sets for the three loci. We retrieved data for *OSPL11* from Harmonizome (see Web Resources), which is a collection of information about genes and proteins from 121 datasets provided by 72 online resources. We also analyzed the protein-protein interaction network of *OSBPL11* using data from the STRING database (see Web Resources). We also examined the genomic context and regulatory annotations of rs2979356 using data from the Haploreg database (see Web Resources). We searched for information on rs2979356 and *OSPL11* in the NHGRI-EBI GWAS catalog, PheGenI, dbGaP, ClinVar, PopHumanScan or in EGA, and Relate to identify their associations with diseases, phenotypes, population-specific variations, clinical significance and evidence of natural selection (see Web Resources for links). We additionally investigated the impact of our lead *OSPL11* variant on risk of severe malaria within the MalariaGEN dataset^21^ and on risk of tuberculosis infection in the International Tuberculosis Host Genetics Consortium^22^(see Web Resources) to determine whether malaria and/or tuberculosis could be responsible for the observed selection pressure. We searched the International Mouse Phenotyping Consortium (IMPC) server for data on the effect of knocking out *Osbpl11* in mice. IMPC conducted a comprehensive panel of phenotyping screens to characterize the *Osbpl11* single-gene knockout mice in comparison to wild-type controls.

### Gene-based analyses

We used the GWAS summary data to calculate gene-based *P*-values for association between genes and 25(OH)D concentrations using MAGMA (v1.08)^39^ gene analysis as implemented in FUMA^54^ using standard settings. In total, 18,912 genes were tested, and significance of *P*-values assessed based on Bonferroni-corrected significant threshold *P*-values of 0.05/18,912=2.644 × 10^-*6*^. We also used MAGMA through the FUMA interface to perform gene set analyses to evaluate the association between 25(OH)D concentrations and biological pathways. We tested a total of 10,678 biological pathways (5,917 gene ontology (GO) terms and 4,761 curated gene sets from the Molecular Signatures Database (MsigDB)^61,62^), and those with *P*-values below a Bonferroni-corrected significance threshold of 1 × 10^-6^ were considered significant.

### Transferability of established 25(OH)D loci

We looked for evidence of transferability of previously reported loci in African diaspora^7^ and European populations^3^ to continental Africans. Variants with *P*-values < 0.05 and the same direction of effect in both European and African populations were considered as transferable. For those variants that were missing or did not show a significant association in our GWAS, we examined all variants with LD r^2^ > 0.3 and within 250 kb of the reported index variant for association with 25(OH)D concentrations. When multiple SNPs from a region were evaluated, the nominal association *P* -values were adjusted for the total number of SNPs within the region using the method of effective degrees of freedom^63,64^. We considered a locus to be replicated if at least one of the tested SNPs had a *P* value of less than the adjusted association *P* value.

### Molecular docking of *OSBPL11* to common oxysterols and vitamin D metabolites

We performed molecular docking analyses using PyRx version 0.9^65^, a graphical user interface for AutoDock Vina^66^. We retrieved the 3D structure of OSBPL11 (PDB format) from AlphaFold Protein Structure Database (see Web Resources). We prepared and optimized the 3D structure for docking by removing any water molecules and generating missing side chains and loop regions using PyRx. Spatial Data File (SDF) format files of common oxysterols (25-hydroxycholesterol, 7-α-hydroxycholesterol, 27-hydroxycholesterol, and 24(S)-hydroxycholesterol), and vitamin D metabolites (namely vitamin D2, vitamin D3, 1,25-dihydroxyvitamin D and 25-hydroxyvitamin D) were retrieved from the PubChem database (see Web Resources). The ligand structures were prepared using PyRx to generate 3D coordinates and protonate the ligands at pH 7.0. The oxysterol binding site of OSBPL11 was defined using the receptor grid generation tool in PyRx with the binding site centered on the box. The docking parameters were set to default values, and 10 docking runs were performed for each ligand. The docking results were analyzed using the PyMOL Molecular Graphics System (Version 2.4) to visualize the protein-ligand interactions and identify the best binding conformation. We used R version 4.0.0 software to plot the lowest binding energies for each oxysterol and vitamin D metabolite for the binding conformation.

### OSBPL11 Alpha Fold modeling and ligand docking

The OSBPL11 Alpha Fold model was downloaded and used without alteration. Ligand docking was carried out using the AutoDock Vina (v. 4.2.6) plugin through UCSF Chimera (v. 1.17.3). Ligands were generated from their CID code and minimized before docking. All components were protonated using Gasteiger. Docking was carried out three times, each with unique seed values; this produced essentially identical poses but with small variations in the resulting score (kcal/mol) values. Figures were made in UCSF Chimera, and the graph was generated using Prism 10 v. 10.2.3.

### OSBPL11 *in vitro* ligand binding assay

Materials: [^3^H]-25-hydroxycholesterol (25[26,27 ^3^H]-OHC) (catalog # 50-353-012) was purchased from Fischer Scientific. Calcitriol (1,25-hydroxyvitamin D) was purchased from MedChemExpress (catalog # HY-113134). Cabot Norit A2 (catalog # DARCO Cas:7440-44-0) charcoal obtained from Sigma Aldrich and Dextran-500 (catalog # D1004) purchased from Spectrum chemicals. Microscint-20 (part.no. 6013621) was purchased from Perkin Elmer. pcDNA3.1 (+)myc/his C was cloned by and purchased from GenScript. Binding assay adapted from established OSBP/ORP *in vitro* binding ^3^H-25-hydroxycholesterol charcoal/dextran binding assay as described^17–19^. OSBPL11cDNA was cloned into pcDNA3.1 (+)myc/his C mammalian expression plasmid, which was transfected into 293T cells with Lipofectamine2000. OSBPL11-myc-His 293T lysate was subjected to a 100,000 g ultracentrifugation, and OSBPL11-myc-His expression verified by protein immunoassay (Supplementary Fig. 11A). For calculation of Ki values in the competition binding curves, a KD of 25 nM for 25-OHC against OSBP was used, based on the measured KD of 25-OHC against OSBPL11^18^.

### Effect of *OSBPL11* lead variant (rs2979356) on cardiometabolic traits in African ancestry populations

In this study, we gathered effect size estimates (beta coefficients and corresponding standard errors) for the lead *OSBPL11* variant, rs2979356, from previously conducted genetic association analyses across multiple African ancestry cohorts. These analyses, carried out by each respective study or consortium (African American Diabetes Mellitus (AADM), UK Biobank, African American Cardiometabolic Disease Knowledge Portal (AMP-CMDKP), Africa Wits-INDEPTH Partnership for Genomic Studies (AWI-Gen), and the Global Lipids Genetics Consortium (GLGC)), focused on cardiometabolic traits such as body composition, lipid profiles, blood pressure, and measures of glucose homeostasis. The AADM study enrolled participants of East and West African ancestry to investigate cardiometabolic diseases. The AWI-Gen study recruited individuals from multiple sub-Saharan African countries (Burkina Faso, Ghana, Kenya, Namibia, South Africa, and Tanzania) to examine genetic and environmental risk factors for cardiometabolic traits. The AMP-CMDKP collates data from diverse African ancestry populations to identify genetic contributions to cardiometabolic diseases. GLGC is a large-scale collaboration focusing on lipid traits in ethnically diverse populations. Finally, the UK Biobank (UKB) African ancestry subset includes participants of African descent within the broader UKB cohort. After confirming consistency in variant reference alleles, each study’s results were extracted in their final, post–quality control form. No additional analyses were performed by the authors; rather, the compiled data were integrated into a single summary table (Table 2), highlighting the associations of rs2979356 with various phenotypes and emphasizing its potential relevance to both lipid metabolism and cardiovascular health in African ancestry populations.

### Web resources (links to web resources that were used in the study)

For H3Africa GWAS pipeline version 3 see https://github.com/h3abionet/h3agwas. For FUMA see https://fuma.ctglab.nl/. All of Us Research Program see https://databrowser.researchallofus.org/. For Sanger Imputation Service see https://imputation.sanger.ac.uk/. Description of Jackson Heart Study see www.jacksonheartstudy.org. For METASOFT see http://genetics.cs.ucla.edu/meta/index.html. For Open Targets Genetics database https://genetics.opentargets.org/. For GTEx portal v8 see https://gtexportal.org/home/. For GTEX Portal see https://gtexportal.org/. For single cell atlas of human and mouse white adipose tissue https://singlecell.broadinstitute.org/single_cell/study/SCP1376/a-single-cell-atlas-of-human-and-mouse-white-adipose-tissue. For AlphaFold Protein Structure Database see https://alphafold.ebi.ac.uk/. For AutoDock Vina https://vina.scripps.edu/. For UCSF Chimera https://www.cgl.ucsf.edu/chimera/. For Ensembl Variant Effect Predictor see https://www.ensembl.org/vep. For Harmonizome see https://maayanlab.cloud/Harmonizome/. For NHGRI-EBI GWAS catalog see https://www.ebi.ac.uk/gwas. For STRING see https://string-db.org/.For ClinVar see https://www.ncbi.nlm.nih.gov/clinvar/. For PheGenI see https://www.ncbi.nlm.nih.gov/gap/phegeni. For dbGaP see https://www.ncbi.nlm.nih.gov/gap/. For data from Accelerating Medicines Partnership in Common Metabolic Diseases Knowledge Portal (AMP-CMDKP) see https://hugeamp.org/.

For EGA see https://ega-archive.org/. For PheWAS see https://gwas.mrcieu.ac.uk/phewas. For HaploReg v4.2 see https://pubs.broadinstitute.org/mammals/haploreg/haploreg.php. For LDpair see https://ldlink.nih.gov/?tab=ldpair. For African Functional Genomics Resource (AFGR) see https://github.com/smontgomlab/AFGR. For JHS eQTL see http://jhsqtl.genetics.unc.edu/eQTL.php. For PubChem database see https://pubchem.ncbi.nlm.nih.gov/. For PhenoScanner database see http://www.phenoscanner.medschl.cam.ac.uk/. For FINEMAP see http://christianbenner.com/. For PolyPhen see http://genetics.bwh.harvard.edu/pph2/. For SIFT see https://sift.bii.a-star.edu.sg/. For dbSNP see https://www.ncbi.nlm.nih.gov/snp/. For PopHumanScan see https://pophumanscan.uab.cat/. For MalariaGEN see https://apps.malariagen.net/. For International Mouse Phenotyping Consortium data see https://www.mousephenotype.org/data/genes/MGI:2146553.

## Data availability

The GWAS summary statistics are available at GWAS catalog (Accession number XXX). The data and analyses scripts underlying this article are available in Harvard Dataverse at XXXXX and applications for data access can be made through the Kilifi Data Governance Committee.

## Acknowledgement

We express our gratitude to our colleagues at the KEMRI-Wellcome Trust Research Programme, specifically Jedidah Mwacharo and Barnes Kitsao, for their assistance in retrieving archived samples. Additionally, we extend our thanks to the teams at the UVRI/MRC Entebbe Mother and Baby Study, the Malaria Vectored Vaccines Consortium (MVVC) in Burkina Faso, and the Respiratory and Meningeal Pathogens Unit (RMPRU) in Johannesburg, South Africa. We would also like to acknowledge the Oxford Genomics Centre at the Wellcome Centre for Human Genetics, which was funded by a Wellcome Trust grant, reference 203141/Z/16/Z, for their role in generating and processing the genotype data for samples from Kenya and The Gambia. MESA and the MESA SHARe project are conducted and supported by the National Heart, Lung, and Blood Institute (NHLBI) in collaboration with MESA investigators. Support for MESA is provided by contracts N01-HC95159, N01-HC-95160, N01-HC-95161, N01-HC-95162, N01-HC-95163, N01-HC-95164, N01-HC-95165, N01-HC95166, N01-HC-95167, N01-HC-95168, N01-HC-95169, UL1-RR-025005, and UL1-TR-000040. Funding support for the Vitamin D dataset was provided by grant HL096875. Funding for SHARe genotyping was provided by NHLBI Contract N02-HL-64278. Genotyping was performed at Affymetrix (Santa Clara, California, USA) and the Broad Institute of Harvard and MIT (Boston, Massachusetts, USA) using the Affymetrix Genome-Wide Human SNP Array 6.0. We thank the International Tuberculosis Host Genetics Consortium (ITHGC) for providing access to GWAS summary statistics. The Jackson Heart Study is supported by Contracts HHSN268201800010I, HHSN268201800011I, HHSN268201800012I, HHSN268201800013I, HHSN268201800014I, HHSN268201800015I from the National Heart Lung and Blood Institute (NHLBI) with additional support from the National Institute of Minority Health and Health Disparities (NIMHD). The views expressed in this manuscript are those of the authors and do not necessarily represent the views of the National Heart, Lung, and Blood Institute; the National Institute of Minority Health and Health Disparities (NIMHD); the National Institutes of Health; or the U.S. Department of Health and Human Services. This study was published with the permission of the Office of the Director of KEMRI.

## Description of Additional Supplementary Files

**Title: Supplementary Dataset 1. FUMA gene-based analysis results**

**Description:** Gene-based analysis results. Analyses were conducted with MAGMA v1.08, using default parameters (SNP-wide mean model) and with the African 1000 Genome Phase3 selected as reference panel. Columns are: GENE, gene Ensembl ID; CHR, chromosome; START/STOP, annotation boundaries of the gene on that chromosome; NSNPs, number of SNPs annotated to the gene that were found in the data and were not excluded based on internal SNP QC; NPARAM, number of relevant parameters used in SNP-wise (mean) model; N, sample size; ZSTAT, the Z-value for the gene, based on its (permutation) P-value (this is what is used as the measure of gene association in the gene-level analyses); P, the gene P-value, using asymptotic sampling distribution (if available); SYMBOL, Gene name.

**Title: Supplementary Dataset 2. MAGMA gene-set analysis output**

**Description:** Gene sets were updated for MsigDB v7.0. Total 15496 gene sets (Curated gene sets: 5500, GO terms: 9996) were tested. Curated gene sets consist of 9 data resources including KEGG, Reactome and BioCarta. GO terms consists of three categories, biological processes (bp), cellular components (cc) and molecular functions (mf). All parameters were set as default (competitive test). Columns are: VARIABLE, name of the gene set, gene covariate or interaction. Names in this column are capped at 30 (by default) characters to keep the output file more readable. FULL_NAME, the full variable name; only included if the variable names exceed the maximum length for the VARIABLE column (and the abbreviate option is not set to file mode); TYPE, denotes the type of variable, either SET or COVAR for normal gene sets and gene covariates provided in the input files; or INTER-SS or INTER-SC, for internally created interaction terms (set by set and set by covariate, respectively); NGENES, the number of genes in the data that are in the set (for gene sets and set by covariate interactions), that are in the interaction set (for set by set interactions), or for which non-missing values were available (for gene covariates); BETA, the regression coefficient of the variable; BETA_STD, the semi-standardized regression coefficient, corresponding to the predicted change in Z-value given a change of one standard deviation in the predictor gene set / gene covariate (i.e.. BETA divided by the variable’s standard deviation). SE, the standard error of the regression coefficient. P, p-value for the parameter / variable.

**Title: Supplementary Dataset 3. FUMA results for annotation of genes to tissue enriched expression.**

**Description:** Tissue expression analysis. MAGMA gene-property test was performed for average gene-expression per category (e.g. tissue type or developmental stage) conditioning on average expression across all categories (one-side). Analysis was conducted with MAGMA v1.06. Columns are: VARIABLE, name of the gene set, gene covariate or interaction; TYPE, the type of variable, either SET or COVAR for normal gene sets and gene covariates provided in the input files; NGENES, the number of genes in the data that are in the set (for gene sets and set by covariate interactions), that are in the interaction set (for set by set interactions), or for which non-missing values were available (for gene covariates); BETA, the regression coefficient of the variable; BETA_STD, semi-standardized regression coefficient, corresponding to the predicted change in Z-value given a change of one standard deviation in the predictor gene set / gene covariate (i.e.. BETA divided by the variable’s standard deviation); SE, standard error of the regression coefficient; P, P-value for the parameter/variable; FULL_NAME: the full variable name; only included if the variable names exceed the maximum length for the VARIABLE column. General parameters and settings for the analysis are included at the top. These are: TOTAL_GENES, the total number of genes included in the analysis; TEST_DIRECTION, the testing direction used for the different types of parameters; CONDITIONED_INTERNAL/RESIDUALIZED/HIDDEN/VARIABLES, the internal covariates and external variables which the analysis was conditioned on.

**Title: Supplementary Dataset 4. GWAS summary statistics and model-averaged posterior summaries for variants predicted to be causal**

**Description:** File, the GWAS summary statistics and model-averaged posterior summaries for each variant one per line. Columns are: RSID, the variant identifiers (rsID number); CHROMOSOME, the chromosome names; POSITION, the base pair positions; ALLELE1, the “first” allele of the variants; ALLELE2, the “second” allele of the variants; MAF, the minor allele frequencies; BETA, the estimated effect sizes as given by GWAS software; SE, the standard errors of effect sizes as given by GWAS software; Z, the z-scores; PROB, the marginal Posterior Inclusion Probabilities (PIP). The PIP for the l^th^ variant is the posterior probability that this variant is causal; LOG10BF, the log10 Bayes factors. The Bayes factor quantifies the evidence that the lth variant is causal with log10 Bayes factors greater than 2 reporting considerable evidence; MEAN, the marginalized shrinkage estimates of the posterior effect size mean for the same allele as in beta; SD, the marginalized shrinkage estimates of the posterior effect size standard deviation; MEAN_INCL, the conditional estimates of the posterior effect size mean for the same allele as in column beta; SD_INCL, the conditional estimates of the posterior effect size standard deviation.

**Title: Supplementary Dataset 5: eQTL Summary Statistics for Multiple Tissues**

**Description:** This dataset contains the eQTL (expression quantitative trait loci) summary statistics for variant rs2979356 across various tissues. Each row represents a unique tissue and provides information about the associated gene, the variant, the effect size, and the significance of the association. Columns: Gencode Id: The unique identifier for each gene from the Gencode database; Gene Symbol, the common symbol or name of the gene; Variant Id, The identifier for the variant, indicating its genomic location and specific alteration; variant Id, The dbSNP identifier for the single nucleotide polymorphism (SNP); P-Value, The statistical significance of the eQTL effect in each tissue. Lower values indicate higher significance; NES (Normalized Effect Size), The magnitude of the eQTL effect. Positive values indicate an increase in gene expression, and negative values indicate a decrease; Tissue, the specific tissue where the eQTL effect was observed.

**Title: Supplementary Dataset 6: Gene Associations for OSBPL11 from the Harmonizome Database**

**Description:** This file contains gene association data for OSBPL11 across various biological datasets, retrieved from the Harmonizome database. Each row represents a specific association between OSBPL11 and a biological trait, condition, or molecular feature. The columns are as follows: ASSOCIATION, the biological trait, condition, or molecular feature associated with OSBPL11 (e.g., tissue types, gene expression profiles, transcription factor binding sites); DATASET, the source dataset from which the association was derived (e.g., Allen Brain Atlas, ENCODE, GTEx, TCGA); THRESHOLD VALUE, the cutoff for determining the significance of the association, with values of 1 or -1 indicating positive or negative associations; STANDARDIZED VALUE, The magnitude of the association between OSBPL11 and the trait, with positive values indicating upregulation and negative values indicating downregulation in the relevant context. Null values indicate cases where no significant data were available.

**Title: Supplementary Dataset 7: PheWAS Summary Statistics for rs2979356 Associated with Various Health Traits.**

**Description:** This file contains PheWAS (Phenome-Wide Association Study) summary statistics for the variant rs2979356, which has been linked to multiple health traits. Each row represents a specific trait association. The columns are as follows: ID: the unique identifier for each trait or study; TRAIT, the health trait associated with rs2979356; POSITION, the base pair position of the variant on chromosome 3; P: the p-value, indicating the statistical significance of the association; SE: the standard error of the effect size estimate; N, the sample size used in each analysis (if available); BETA, the estimated effect size of the variant on the trait; CHR, the chromosome number where rs2979356 is located; EA, the effect allele of rs2979356; NEA, the non-effect allele of rs2979356; EAF, the effect allele frequency in the population.

